# Network Analysis of Rare Single-Nucleotide Polymorphisms Suggests a Central Role for Type 17 Immune Responses in Trauma-induced, Genotypically Associated Hypo-Inflammation and Critical Illness

**DOI:** 10.1101/2025.01.28.25321184

**Authors:** Fayten El-Dehaibi, Ruben Zamora, Jinling Yin, Timothy R. Billiar, Yoram Vodovotz

## Abstract

Critical illness stemming from severe traumatic injury is a leading cause of morbidity and mortality worldwide, involves the dysfunction of multiple organ systems, and is driven at least in part by dysregulated inflammation that involves type 17 immunity. We and others have demonstrated a genetic predisposition to adverse critical illness outcomes associated with single-nucleotide polymorphisms (SNPs) in novel loci distinct from those which impact type 17 immune responses yet acting in concert with those responses. We have recently developed *SNPScanner*, an algorithm that enables rapid scanning through a large SNP dataset and associated inflammation biomarkers and clinical data, and in the present study leveraged this algorithm in concert with existing bioinformatics tools to define networks of interaction among rare SNPs (defined operationally as being present in 5-10% of patients due to the study cohort size) assessed in critically ill trauma patients. RAR-related orphan receptor A (RORA), a transcription factor central to the differentiation of Th17 cells, was inferred as a hub gene via its SNP rs4774381; stratification of trauma patients based on homozygous genotype at this SNP pointed to distinct organ dysfunction trajectories associated with network-defined hypo-inflammation. Further, the SNP rs11919443 in the non-hub *TM4SF19* gene distinguished genotype-associated systemic IL-17A trajectories, and associated hypo-inflammation with adverse outcomes. In contrast, no Th17-related hub genes were identified in a control group of SNPs associated algorithmically with adverse clinical outcomes but with genotypically indistinct systemic inflammatory responses; the main inferred hub gene in this control group was solute carrier family 2, facilitated glucose transporter member 1 (*SLC2A1*)/glucose transporter 1 (GLUT1). Secondary analysis showed several statistically significant differences in circulating inflammatory mediators not including IL-17A, and also associated network-based hypo-inflammation with adverse outcomes in this control group. This study thus extends our prior work aimed at defining genetic predisposition to dysregulated inflammation and pathophysiology in the context of critical illness, and points to a crucial role for type 17 immune responses.

## 1. INTRODUCTION

Trauma-induced critical illness, in which organ dysfunction is associated with dysregulated inflammation (i.e., an abnormal or imbalanced immune response with detrimental effects to the body), represents a major global health burden (1–3). Despite similar injury profiles and demographics, and the use of standardized protocols, critically ill trauma patients exhibit heterogeneous outcomes (4). We and others have proposed that genetic variation among patients may account for some of this heterogeneity (5). Previous studies have identified distinct dynamic inflammatory networks after severe trauma as a function of demographics (e.g., age) (6), injury timing (7) and characteristics (8–11), or clinical outcomes (12, 13). We have also characterized endotypes of blunt trauma patients based on early inflammatory patterns (14, 15), multiple organ dysfunction severity (16), or multi-‘omics patterns (17, 18). A key immune pathway that emerged from these studies involves interleukin-17A (IL-17A), suggesting a novel, early role for type 17 immune responses in trauma-associated critical illness (13, 15, 19–21).

Our studies have also highlighted a role for genetic polymorphisms in the disordered systemic inflammation that is characteristic of critically ill trauma patients (19, 22–25). These studies, particularly those showing that specific single-nucleotide polymorphisms (SNPs) are linked to sub-acute mortality in trauma patients with early divergent inflammatory trajectories associated with the Multiple Organ Dysfunction Syndrome (MODS) (13, 24), imply that genetic factors may influence the variability in patient outcomes and acute inflammation in critically ill trauma patients.

We have previously developed novel computational approaches to aid in the search for novel, yet common (defined as present in 20% or more of the patients in our study cohort) SNPs associated with altered trajectories of systemic inflammation and subsequent clinical outcomes (24), and have combined this discovery program with existing computational tools that facilitate the discovery of SNPs in networks of genes acting in concert (25). In the present study, we leveraged these two approaches to scan a dataset of >500,000 SNPs for networks of both rare and common SNPs associated with adverse outcomes in critically ill trauma patients. In these analyses, we focused on defining hub genes and assessing clinical outcomes and inflammatory profiles in patient subgroups stratified based on genotypes at those hub genes. Our results suggest a key role for Th17 differentiation and associate this with systemic hypo-inflammation.

## 2. MATERIALS AND METHODS

### 2.1 Trauma patient cohort

Studies were carried out using a cohort of 380 blunt trauma survivors with roughly equivalent distributions of homozygous and alternative homozygous genotypes at any of >500,000 SNPs described previously (24). In brief, patients were enrolled by screening after presentation to the emergency department of the UPMC Presbyterian hospital (a Level 1 trauma center) between May 5, 2004, to May 2, 2012. Patients eligible for enrollment in the study were at least 18 years of age, admitted to the intensive care unit (ICU) after being resuscitated, and, per treating physician, were expected to live more than 24h. Exclusion criteria were isolated head injury, pregnancy, and penetrating trauma. Informed consent was obtained from each patient or next of kin as per the University of Pittsburgh Institutional Review Board (IRB; Protocol No. MOD08010232-19 / PRO08010232) and in accordance with the Declaration of Helsinki. Blood samples for DNA and inflammatory biomarker analysis were obtained upon admission to the trauma bay, at first 24 h of hospitalization, and daily thereafter for 7 days.

Clinical data, including injury severity score (ISS), abbreviated injury scale (AIS) score, Marshall Multiple Organ Dysfunction (MOD) score, admission Shock Index, ICU LOS, hospital LOS, and days on mechanical ventilation were collected from the hospital inpatient electronic and trauma registry database. ISS (26) and AIS (27) were calculated for each patient by a single trauma surgeon after attending radiology evaluations were finalized. The ISS is based on an anatomical scoring system that provides an overall score for patients with multiple injuries. Each injury is assigned an AIS score, allocated to one of six body regions: head, face, chest, abdomen, extremities (including pelvis), and external. As an index of organ dysfunction, the MOD score (28) ranging from 0 to 24 was calculated. In brief, six variables were obtained from the electronic trauma data registry including (a) the respiratory system (PO_2_/FIO_2_ ratio); (b) the renal system (serum creatinine concentration); (c) the hepatic system (serum bilirubin concentration); (d) the hematologic system (platelet count); (e) the central nervous system (Glasgow Coma Scale); and (f) the cardiovascular system- the pressure-adjusted heart rate (PAR).

The overall characteristics of the patients in the trauma cohort were as follows (mean ± SD): (ISS) = 19.0 ± 10.1 [min: 1; max: 54]; mean age: 49.3 ± 19.2 years (min: 18y; max: 90y); 119 women, 261 men; 96.84% whites, 2.63% blacks and 0.53% others.

### 2.2 Genomic analyses

For the blunt trauma patient cohorts, DNA was prepared from whole blood samples and analyzed using Illumina® arrays as described previously (19, 22, 23). Whole blood samples were collected into EDTA-treated tubes and DNA was extracted using the QIAamp® DNA Blood Midi Kit (QIAGEN, Valencia, CA) as per manufacturer’s specifications. Single nucleotide polymorphism genotyping was performed with 200 ng of genomic DNA input using the Human Core Exome-24 v1.1 BeadChip (Illumina, San Diego, CA) following the manufacturer’s Infinium® HTS Assay protocol. This array interrogated 551,839 SNPs, including both SNPs previously implicated in genome-wide association studies as well as additional coding and non-coding SNPs spaced along the human genome. Briefly, DNA was denatured in 0.1N NaOH and neutralized prior to isothermal amplification. Amplified DNA was fragmented and then hybridized to locus-specific 50mers that make up the array for 16-24 h with rocking at 48°C. After removal of unbound or non-specifically annealed DNA, single base extension of the 50mer oligonucleotides was performed with labeled nucleotides, which were scanned using an Illumina iScan with autoloader 2.x. Data analysis was performed using Illumina Genome Studio 2.0.

### 2.3 Assays of inflammation biomarkers

Whole-blood samples were drawn in EDTA-treated tubes, which were kept on ice and centrifuged to obtain plasma and then stored at -80°C until assays were performed. The Luminex™ 100 IS analyzer (Luminex, Austin, TX) and Human Cytokine/Chemokine MILLIPLEX™ Panel kit (Millipore Corporation, Billerica, MA) were used to measure plasma levels of Eotaxin (CCL11), interleukin (IL)-1β, IL-1 receptor antagonist (IL-1RA), IL-2, soluble IL-2 receptor-α (sIL-2Rα), IL-4, IL-5, IL-6, IL-7, IL-8 (CCL8), IL-10, IL-13, IL-15, IL-17A, interferon (IFN)-α, IFN-γ, IFN-γ inducible protein (IP)-10 (CXCL10), monokine induced by gamma interferon (MIG; CXCL9), macrophage inflammatory protein (MIP)-1α (CCL3), MIP-1β (CCL4), monocyte chemotactic protein (MCP)-1 (CCL2), granulocyte-macrophage colony stimulating factor (GM-CSF) and tumor necrosis factor alpha (TNF-α). Human Th17 MILLIPLEX™ Panel kit (Millipore Corporation, Billerica, MA) was used to measure IL-9, IL-21, IL-22, IL-23, IL-17E/25, and IL-33. NO2-/NO3 levels were measured by a Griess Reagent colorimetric assay (Cayman Chemical, Ann Arbor, MI). IL-1 receptor-like 1/ suppression of tumorigenicity-2 (ST2) was measured by a sandwich ELISA assay (R&D Systems, Minneapolis, MN). All cytokine/chemokine concentrations are given in pg/ml; NO2-/NO3-concentrations are in µM. Experimental data are shown as mean ± SEM.

### 2.4 FLAME and associated bioinformatics analyses

Functional Enrichment Analysis was performed using the web application FLAME (v2.0) (29). Two lists of SNPs— the first associated with trajectories of systemic inflammation typical that did not differ based on genotype (which served as controls) vs. SNPs associated with genotype-specific differential systemic inflammatory responses as determined in our prior study (24) —were separately entered into the SNP tab of the Input page (Table 1). To reduce potential artifacts in this analysis, 15 SNPs associated with 12 or more genotype-specific differential systemic inflammatory responses as determined in our prior study (22) were selected out of 7482 SNPs which met the initial criterion of having at least one statistically different biomarker difference as a function of SNP genotype in our original study (22). In the control group for evenly distributed genotypes, SNPs were extracted from the group of 567 SNPs with zero significantly different mediators as a function of genotype at each given SNP (described initially in (22). To decrease the likelihood of artifactual associations among SNPs and to obtain meaningful information about associations of SNPs with inflammation, we imposed an additional condition of having at least 3-4 significant differences in clinical outcomes as a function of SNP genotype which is to find SNPs while having no significant differences in inflammation biomarkers. This led to down-selection from 567 to 16 SNPs.

**Table 1.** ‘Rare’ SNPs entered into FLAME, their number of statistically significant inflammatory biomolecules found by *SNPScanner*, and their corresponding gene/entry found by FLAME (if applicable). (A) SNPs with at least two statistically distinct (p < 0.00001) biomolecular differences between AA and BB according to MWU. (B) SNPs with zero statistically distinct (p < 0.05) biomolecular differences.

| A. ‘Rare’ Inflammatory Disparity SNPs |  |  |
| --- | --- | --- |
| SNP | Number of Distinct Biomolecules<br>($p < 0.00001$) | Gene/Entry |
| rs10489239 | 2 | ENSG00000231424 |
| rs6603352 | 2 | KLHL13 |
| rs4774381 | 2 | RORA |
| rs11782445 | 2 | --- |
| rs7782688 | 2 | --- |
| rs1010779 | 2 | --- |
| rs6603357 | 2 | KLHL13 |
| rs12009735 | 2 | KLHL13 |
| rs6726331 | 2 | LINC01807 |
| rs7258387 | 2 | ENSG00000269110 |
| rs261537 | 2 | UBE2D2 |
| rs13007274 | 2 | FSHR, ENSG00000282890 |
| rs13253492 | 2 | CSMD1 |
| rs7918717 | 2 | RPP38, NMT2 |
| rs11919443 | 2 | TM4SF19, TM4SF19-AS1,<br>TM4SF19-DYNLT2B |
| B. ‘Rare’ Control SNPs |  |  |
| SNP | Number of Distinct Biomolecules<br>($p < 0.05$) | Gene/Entry |
| rs1409117 | 0 | IQSEC2 |
| rs2388252 | 0 | ENSG00000235281 |
| rs12450000 | 0 | --- |
| rs4144013 | 0 | PRRG1, ENSG00000250349 |
| rs6609469 | 0 | --- |
| rs9558839 | 0 | --- |
| rs7609779 | 0 | ABI3BP |
| rs767478 | 0 | --- |
| rs841852 | 0 | SLC2A1 |
| rs7514892 | 0 | RGS21, ENSG00000285280 |

**Table 2.** Clinical summary of patients by genotype for rs4774381 (A), rs11919443 (B), rs7918717 (C), rs261537 (D), and rs841852 (E). Parameters marked with an asterisk (*) have five patients with no recorded Shock Index value and four outliers observed in the available patients’ data.

| A. rs4774381 Genotypes |  |  |  |
| --- | --- | --- | --- |
|  | AA | AB | BB |
| Patient Count | 223 | 136 | 21 |
| Age | 50.55 ± 19.08<br>(min: 18; max: 90) | 46.26 ± 18.70<br>(min: 18; max: 87) | 54.90 ± 20.39<br>(min: 21; max: 85) |
| Sex |  |  |  |
| Male (%) | 154 (69.06%) | 91 (66.91%) | 16 (76.19%) |
| Female (%) | 69 (30.94%) | 45 (33.09%) | 5 (23.81%) |
| Ethnicity |  |  |  |
| White (%) | 220 (98.65%) | 130 (95.59%) | 18 (85.71%) |
| African American (%) | 1 (0.45%) | 6 (4.41%) | 3 (14.29%) |
| Asian (%) | 2 (0.90%) | 0 | 0 |
| ISS | 19.65 ± 10.32<br>(min: 1; max: 54) | 18.26 ± 10.15<br>(min: 1; max: 50) | 17.38 ± 7.38<br>(min: 5; max: 30) |
| ICU Length of Stay (days) | 6.51 ± 7.03<br>(min: 1; max: 36) | 5.64 ± 7.10<br>(min: 1; max: 36) | 7.62 ± 8.81<br>(min: 1; max: 35) |
| Total Hospital Length of Stay (days) | 11.86 ± 9.37<br>(min: 1; max: 48) | 11.44 ± 9.73<br>(min: 2; max: 50) | 13.71 ± 8.11<br>(min: 2; max: 35) |
| Days on Mechanical Ventilation | 3.26 ± 6.17<br>(min: 0; max: 34) | 2.50 ± 5.74<br>(min: 0; max: 37) | 3.90 ± 6.39<br>(min: 0; max: 26) |
| Shock Index* | 0.85 ± 0.70<br>(min: 0.07; max: 8.67) | 0.84 ± 0.85<br>(min: 0.29; max: 9.00) | 0.75 ± 0.24<br>(min: 0.26; max: 1.42) |
| B. rs11919443 Genotypes |  |  |  |
|  | AA | AB | BB |
| Patient Count | 20 | 111 | 249 |
| Age | 57.95 ± 21.10<br>(min: 23; max: 90) | 46.04 ± 17.81<br>(min: 18; max: 87) | 49.99 ± 19.31<br>(min: 18; max: 89) |
| Sex |  |  |  |
| Male (%) | 14 (70%) | 88 (79.28%) | 159 (63.86%) |
| Female (%) | 6 (30%) | 23 (20.72%) | 90 (36.14%) |
| Ethnicity |  |  |  |
| White (%) | 18 (90%) | 108 (97.30%) | 242 (97.19%) |
| African American (%) | 1 (5%) | 2 (1.80%) | 7 (2.81%) |
| Asian (%) | 1 (5%) | 1 (0.90%) | 0 |
| ISS | 18.35 ± 10.23<br>(min: 5; max: 48) | 20.06 ± 10.91<br>(min: 1; max: 54) | 18.62 ± 9.75<br>(min: 1; max: 50) |
| ICU Length of Stay (days) | 9.85 ± 9.02<br>(min: 1; max: 33) | 6.57 ± 7.91<br>(min: 1; max: 36) | 5.83 ± 6.56<br>(min: 1; max: 36) |
| Total Hospital Length of Stay (days) | 16.40 ± 13.04<br>(min: 2; max: 48) | 12.63 ± 9.54<br>(min: 1; max: 45) | 11.08 ± 8.92<br>(min: 2; max: 50) |
| Days on Mechanical Ventilation | 4.40 ± 4.47<br>(min: 0; max: 14) | 3.50 ± 7.54<br>(min: 0; max: 37) | 2.70 ± 5.31<br>(min: 0; max: 28) |
| Shock Index* | 0.68 ± 0.15<br>(min: 0.49; max: 1.04) | 0.87 ± 0.81<br>(min: 0.07; max: 8.67) | 0.84 ± 0.74<br>(min: 0.30; max: 9.00) |
| C. rs7918717 Genotypes |  |  |  |
|  | AA | AB | BB |
| Patient Count | 242 | 117 | 21 |
| Age | 49.78 ± 19.15<br>(min: 18; max: 90) | 46.76 ± 18.79<br>(min: 18; max: 86) | 57.10 ± 19.06<br>(min: 20; max: 83) |
| Sex |  |  |  |
| Male (%) | 168 (69.42%) | 78 (66.67%) | 15 (71.43%) |
| Female (%) | 74 (30.58%) | 29 (24.79%) | 6 (28.57%) |
| Ethnicity |  |  |  |
| White (%) | 239 (98.76%) | 114 (97.44%) | 15 (71.43%) |
| African American (%) | 1 (0.41%) | 3 (2.56%) | 6 (28.57%) |
| Asian (%) | 2 (0.82%) | 0 | 0 |
| ISS | 18.70 ± 10.00<br>(min: 1; max: 50) | 19.40 ± 10.35<br>(min: 1; max: 54) | 20.71 ± 10.48<br>(min: 4; max: 50) |
| ICU Length of Stay (days) | 5.73 ± 6.69<br>(min: 1; max: 33) | 7.00 ± 7.72<br>(min: 1; max: 36) | 8.19 ± 8.70<br>(min: 1; max: 35) |
| Total Hospital Length of Stay (days) | 11.15 ± 8.84<br>(min: 1; max: 50) | 13.30 ± 10.67<br>(min: 2; max: 46) | 11.19 ± 7.95<br>(min: 2; max: 35) |
| Days on Mechanical Ventilation | 2.59 ± 5.28<br>(min: 0; max: 27) | 3.74 ± 7.41<br>(min: 0; max: 37) | 3.95 ± 5.40<br>(min: 0; max: 17) |
| Shock Index* | 0.82 ± 0.75<br>(min: 0.07; max: 9.00) | 0.89 ± 0.79<br>(min: 0.38; max: 8.67) | 0.73 ± 0.22<br>(min: 0.41; max: 1.22) |
| D. rs261537 Genotypes |  |  |  |

|  | AA | AB | BB |
| --- | --- | --- | --- |
| Patient Count | 223 | 136 | 21 |
| Age | 49.47 ± 19.19<br>(min: 18; max: 86) | 48.20 ± 19.11<br>(min: 18; max: 90) | 53.86 ± 18.72<br>(min: 23; max: 85) |
| Sex |  |  |  |
| Male (%) | 154 (69.06%) | 91 (66.91%) | 16 (76.19%) |
| Female (%) | 69 (30.94%) | 45 (33.09%) | 5 (23.81%) |
| Ethnicity |  |  |  |
| White (%) | 219 (98.2%) | 130 (95.59%) | 19 (90.48%) |
| African American (%) | 2 (0.90%) | 6 (4.41%) | 2 (9.52%) |
| Asian (%) | 2 (0.90%) | 0 | 0 |
| ISS | 18.53 ± 10.23<br>(min: 1; max: 54) | 20.15 ± 10.20<br>(min: 4; max: 50) | 17.86 ± 8.28<br>(min: 5; max: 42) |
| ICU Length of Stay (days) | 6.24 ± 7.18<br>(min: 1; max: 36) | 6.62 ± 7.41<br>(min: 1; max: 35) | 4.14 ± 5.05<br>(min: 1; max: 24) |
| Total Hospital Length of Stay (days) | 11.97 ± 10.00<br>(min: 2; max: 50) | 11.76 ± 8.79<br>(min: 1; max: 45) | 10.48 ± 7.15<br>(min: 2; max: 33) |
| Days on Mechanical Ventilation | 2.91 ± 5.80<br>(min: 0; max: 34) | 3.43 ± 6.66<br>(min: 0; max: 37) | 1.67 ± 3.68<br>(min: 0; max: 17) |
| Shock Index* | 0.86 ± 0.70<br>(min: 0.30; max: 9.00) | 0.83 ± 0.85<br>(min: 0.07; max: 8.67) | 0.68 ± 0.19<br>(min: 0.30; max: 0.99) |
| E. rs841852 Genotypes |  |  |  |
|  | AA | AB | BB |
| Patient Count | 21 | 109 | 250 |
| Age | 57.90 ± 15.83<br>(min: 23; max: 85) | 49.59 ± 19.25<br>(min: 18; max: 87) | 48.38 ± 19.22<br>(min: 18; max: 90) |
| Sex |  |  |  |
| Male (%) | 12 (57.14%) | 80 (73.39%) | 169 (67.60%) |
| Female (%) | 9 (42.86%) | 29 (26.61%) | 81 (32.40%) |
| Ethnicity |  |  |  |
| White (%) | 19 (90.48%) | 104 (95.41%) | 245 (98.00%) |
| African American (%) | 2 (9.52%) | 5 (4.59%) | 3 (1.20%) |
| Asian (%) | 0 | 0 | 2 (0.80%) |
| ISS | 15.86 ± 7.83<br>(min: 1; max: 30) | 17.93 ± 10.23<br>(min: 1; max: 50) | 19.77 ± 10.19<br>(min: 1; max: 54) |
| ICU Length of Stay | 3.86 ± 7.19 | 7.12 ± 7.86 | 6.08 ± 6.81 |
| (days) | (min: 1; max: 35) | (min: 1; max: 33) | (min: 1; max: 36) |
| Total Hospital Length of Stay (days) | 8.43 ± 7.20<br>(min: 2; max: 35) | 12.96 ± 10.24<br>(min: 2; max: 48) | 11.60 ± 9.17<br>(min: 1; max: 50) |
| Days on Mechanical Ventilation | 1.00 ± 2.58<br>(min: 0; max: 12) | 3.58 ± 6.31<br>(min: 0; max: 31) | 2.95 ± 6.10<br>(min: 0; max: 37) |
| Shock Index* | 1.02 ± 1.80<br>(min: 0.07; max: 9.00) | 0.88 ± 0.94<br>(min: 0.42; max: 8.67) | 0.81 ± 0.42<br>(min: 0.26; max: 5.4) |

To examine less-common SNPs (referred to as ‘rare’ for our purposes) we performed the process described to extract differential inflammatory SNPs from the human trauma group but used a 5–10% to 90–95% ratio of homozygous prevalence (minimum 20 patients) to select 7212 SNPs from the original >500,000. The previous method was applied to run Mann-Whitney U analysis on the same 30 inflammatory biomarkers on patients from each group. Given the population size disparity between the homozygotes of the rare SNP set, we sought to reduce the likelihood of artifactual results by considering p-values less than 10-5 as significant, as opposed to 0.05 used earlier. This stringency provided 131 SNPs associated with at least one statistically significant biomarker, and 15 SNPs with up to two. The omission of X-linked SNPs yielded 12 rare somatic SNPs with at least two significantly different inflammatory biomarkers. The rare SNP subset was also checked for control SNPs in the same manner described for the evenly distributed SNPs. Screening clinical outcomes with Mann-Whitney U reduced 433 rare SNPs with zero significantly different mediators (p < 0.05) down to 10 with at least 3-4 statistically different outcomes.

Each list of corresponding genes found was separately inputted into FLAME’s Functional Enrichment page for whole genome search, with the enrichment tools set to gProfiler (30) and enrichR (31) and the significance threshold set to 0.05. The following data sources were selected: Molecular Function, Cellular Component and Biological Function from the gene ontology knowledgebase (GO)(32), and the Kyoto Encyclopedia of Genes and Genomics (KEGG) (33).

### 2.5 Statistical analyses

Kruskal-Wallis Analysis of Variance (ANOVA) on Ranks, One-Way and Two-Way ANOVAs were conducted to analyze the differences in ISS, AIS, Marshall MODScore, and time-dependent changes in plasma biomarkers in AA and BB patients using SigmaPlot (Systat Software, San Jose, CA) as appropriate. Two-Way ANOVA was followed by the Holm-Sidak *post hoc* test to isolate which group(s) differ from the others. Chi-square analysis of the proportions of observations for patients with rs10404939 or rs11117564 genotype was performed using SigmaPlot (Systat Software, San Jose, CA).

### 2.6 Network analysis

Dynamic Network Analysis (DyNA) was carried out to define, in a granular fashion, the central inflammatory network nodes as a function of both time and patient subgroup. DyNA networks were created over six consecutive time periods (D1-D2, D2-D3, D3-D4, D4-D5, D5-D6, D6-D7) using MATLAB® software as described previously(34–36). Connections, defined as the number of inflammatory biomarkers that were positively or negatively correlated across time intervals, were created if the Pearson correlation coefficient between any two nodes (biomarkers) at the same time-interval was greater or equal to a threshold of an absolute value of 0.95, as appropriate. The network complexity for each time-interval was calculated using the following formula: Sum (N_1_+ N_2_+…+ Nn)/(n − 1), where N represents the number of connections for each biomarker and n is the total number of biomarkers analyzed.

### 2.7 Th17 Cell Subset Inference/Correlation Analysis

Spearman’s correlation carried out to measure the strength of the association between the Luminex™ data for two different mediators (IL-17A and each of the following: GM-CSF, IL-10, TNF-α, and IL-22) was generated using MATLAB® software (Natick, MA).

## 3. RESULTS

### 3.1 Bioinformatic analysis defines networks of interaction among rare SNPs in blunt trauma patients and implicates Th17 differentiation and systemic hypo-inflammation

We have previously searched for common SNPs (defined operationally as being present in 20% or more of our study cohort) that might be both unlikely and associated with dysregulated inflammation and adverse clinical outcomes in the context of critical illness, and found one such SNP in the *LYPD4* gene (out of >500,000 assessed SNPs) using a Python-based algorithm we term *SNPScanner* (24).

In the present study, we sought to study less common SNPs and focused on SNPs present in 5-10% of the population to allow for a sufficient number of patients in the various comparison groups. Herein, we refer to these SNPs as “rare” to distinguish them from the “common” *LYPD4* SNP identified previously (24), acknowledging that most SNPs are found in between 0.5 and 5% of the global population, and SNPs found in a greater proportion of individuals are unlikely (37). Our search yielded 12 somatic SNPs associated with two highly statistically significantly different inflammation biomarkers (based on the initial Mann-Whitney U-test performed in the *SNPScanner* algorithm), with a p-value < 0.00001 between subgroups of patients homozygous for the main allele at a given SNP (referred to as AA) vs. the alternative homozygous allele at that SNP (referred to as BB). We then carried out Functional Enrichment Analysis using the web application FLAME (29). When the *SNPScanner* results were entered into FLAME, one of these SNPs, three major and one minor hub gene were identified.

One major hub gene, with three connections, was identified via the SNP rs4774381 in the gene encoding RAR-related orphan receptor A (RORA), which is involved in Th17 differentiation (38) (Figure 1). When segregating trauma patients based on AA vs. BB genotype at this SNP, statistically significant differences were noted in circulating IL-17A along with IFN-γ, IL-1RA, IL-5, IL-7, IL-10, MIG/CXCL9, sIL-2rα, and sST2 (Figure 2A). This was associated with statistically significant differences in MODS trajectories (Figure 2B) but not in admission Shock Index between AA and BB (p=0.799). Paradoxically, rs4774381^AA^ exhibited significantly (p < 0.001) lower Marshall MODScores as compared to rs4774381^BB^ patients despite the elevated systemic inflammation in rs4774381^AA^ vs. rs4774381^BB^ patients (Figure 2B). This finding raised the possibility that pathological hypo-inflammation might be associated with adverse outcomes, as suggested in our prior work (15). In support of this hypothesis, we note the early elevation of systemic IL-10, a key anti-inflammatory cytokine, in rs4774381^BB^ patients as compared to rs4774381^AA^ patients.

**Figure 1.**
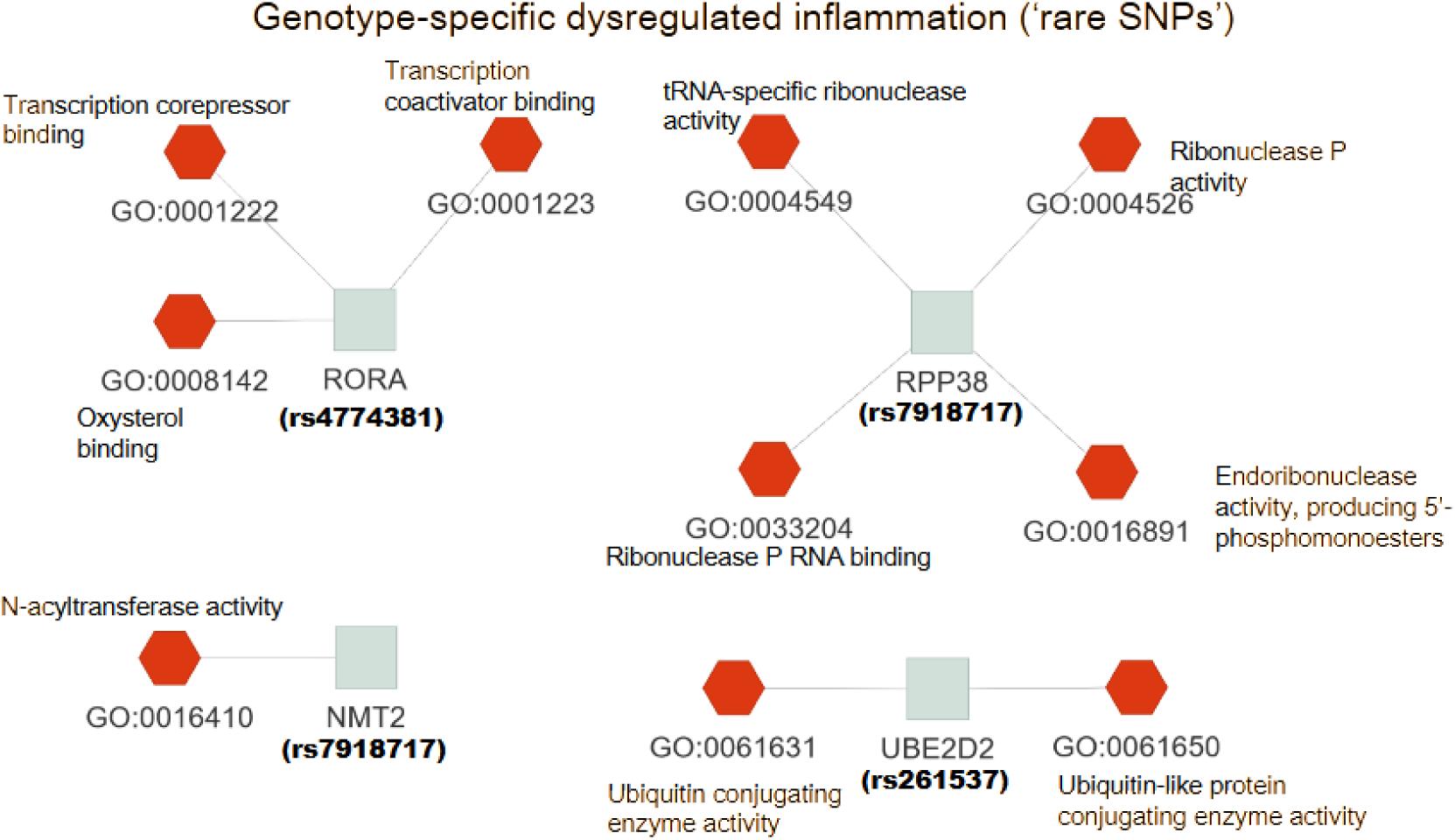
FLAME tree plot describing molecular function connectivity of ‘rare’ SNPs implicated in inflammatory biomolecules disparity by *SNPScanner*. These results were found in the Gene Ontology knowledgebase with the web tool *enrichR*. SNP rs4774381 corresponds to the gene for RORA, which is a component of the mTORc1 complex in a Th17 differentiation pathway.

**Figure 2.**
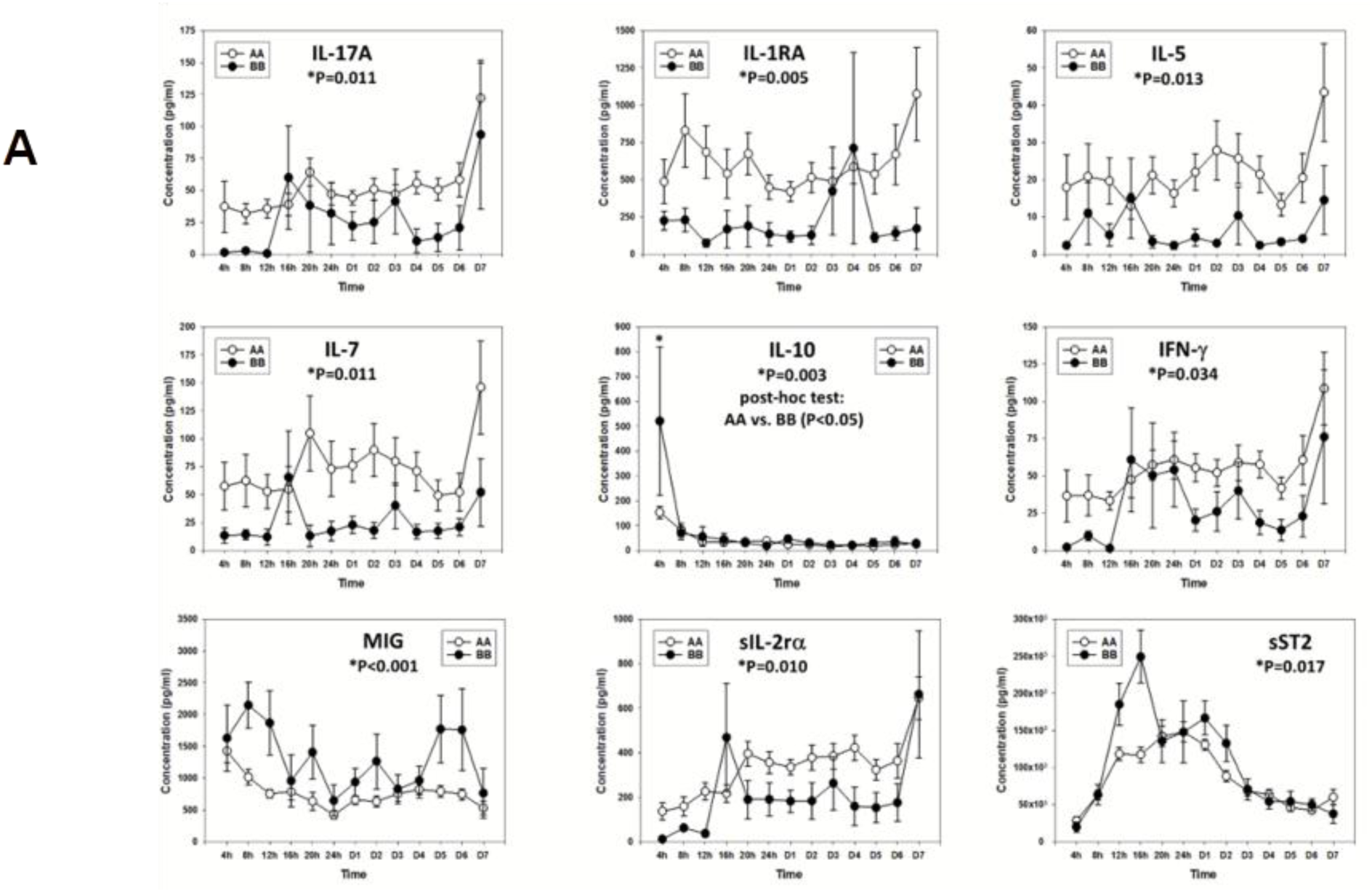
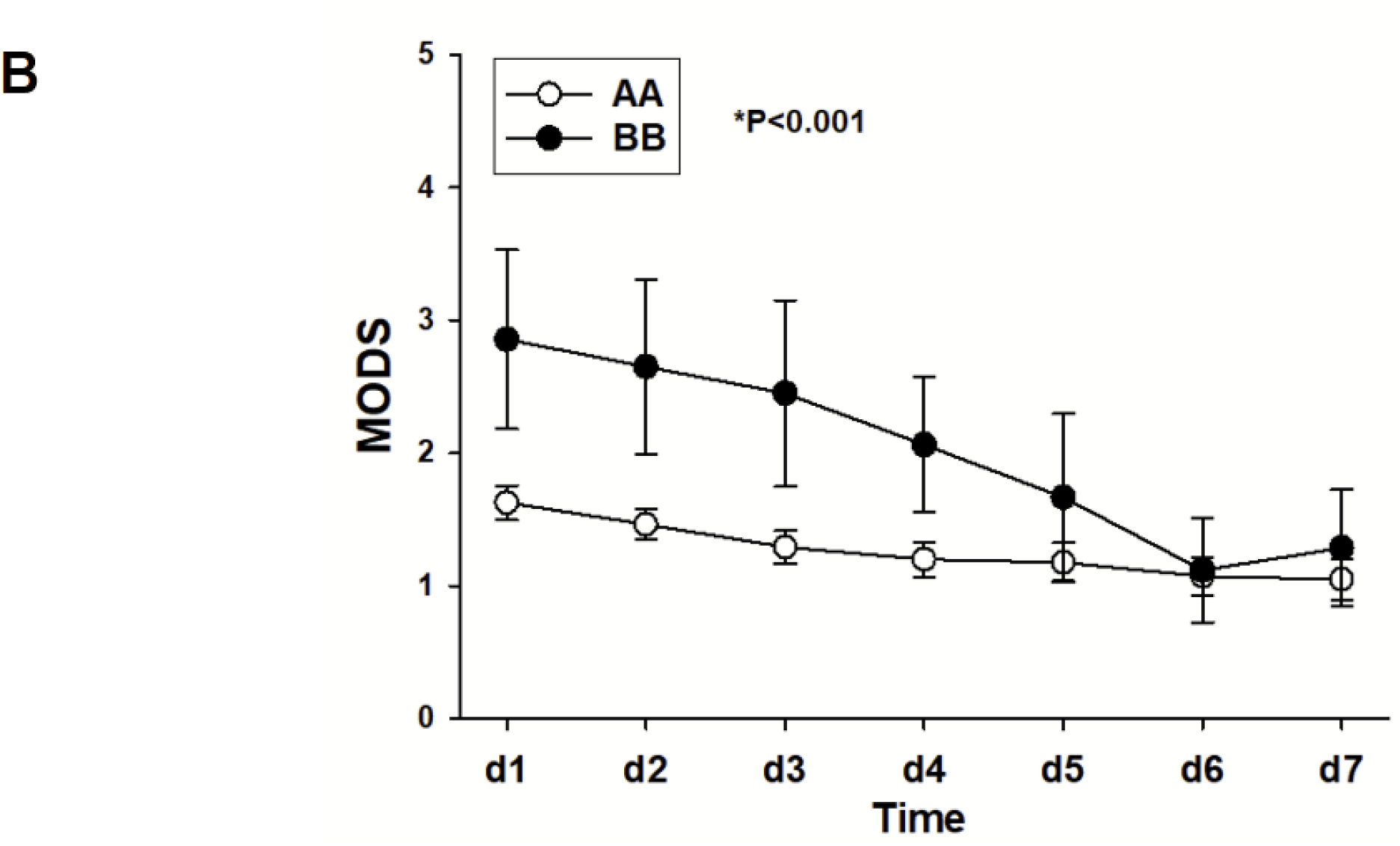
Distinct trajectories of circulating inflammatory mediator mediators as a function of homozygous genotype at rs4774381. **(A)** Statistically distinct (*p < 0.05 by Two-Way ANOVA) circulating inflammatory biomarker concentrations from 4 hours to 7 days past admission between patients with rs4774381^AA^ and rs4774381^BB^. **(B)** Marshall MODS seven-day time course between rs4774381^AA^ and rs4774381^BB^ patients (*p < 0.001 by Two-Way ANOVA).

We next utilized Dynamic Network Analysis (DyNA) (39) to define networks of interaction among all systemic inflammatory mediators in rs4774381^AA^ vs. rs4774381^BB^ patients (Figure 3). This analysis demonstrated striking differences in network patterns between these two patient subgroups. Specifically, the dynamic networks of systemic inflammation inferred in rs4774381^AA^ (Figure 3A, 3C) were much sparser at early time points post-hospitalization than those inferred in s4774381^BB^ patients (Figure 3B, 3C). In addition, both rs4774381^AA^ and rs4774381^BB^ patients exhibited oscillating systemic inflammation networks over the first seven days following hospitalization, with the amplitude of oscillation appearing to manifest a “phase shift” when comparing the two patient subgroups (Figure 3C).

**Figure 3.**
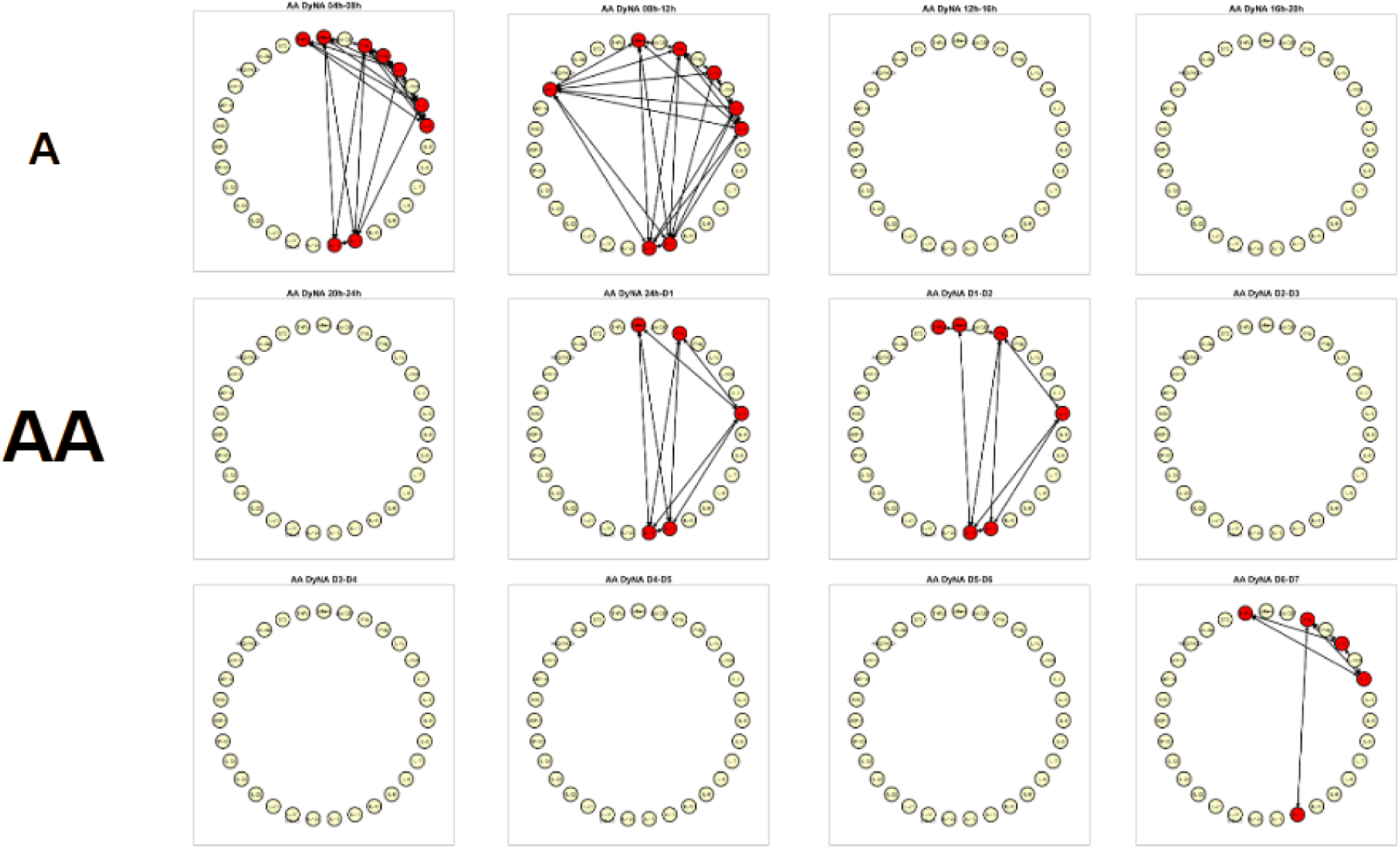
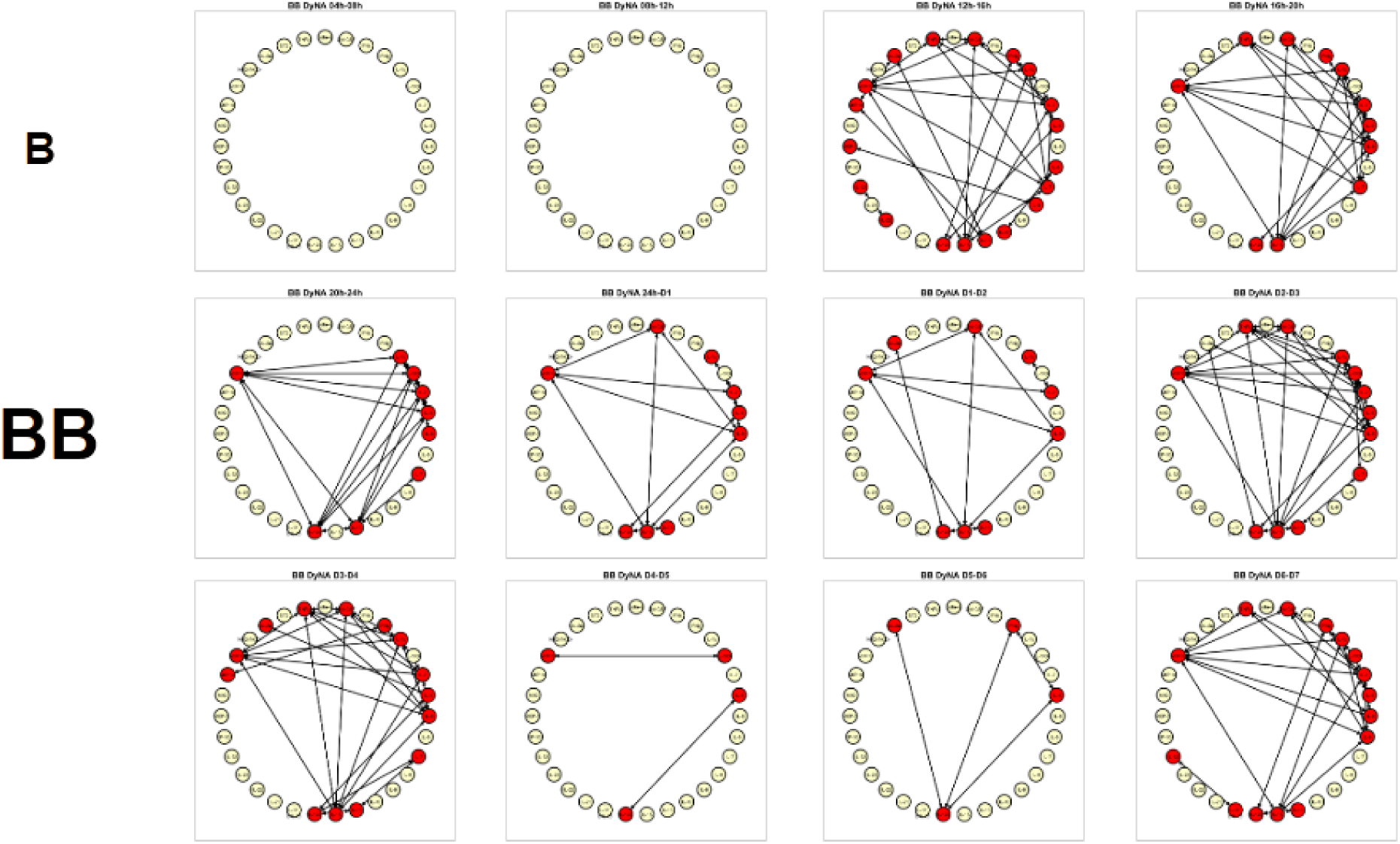
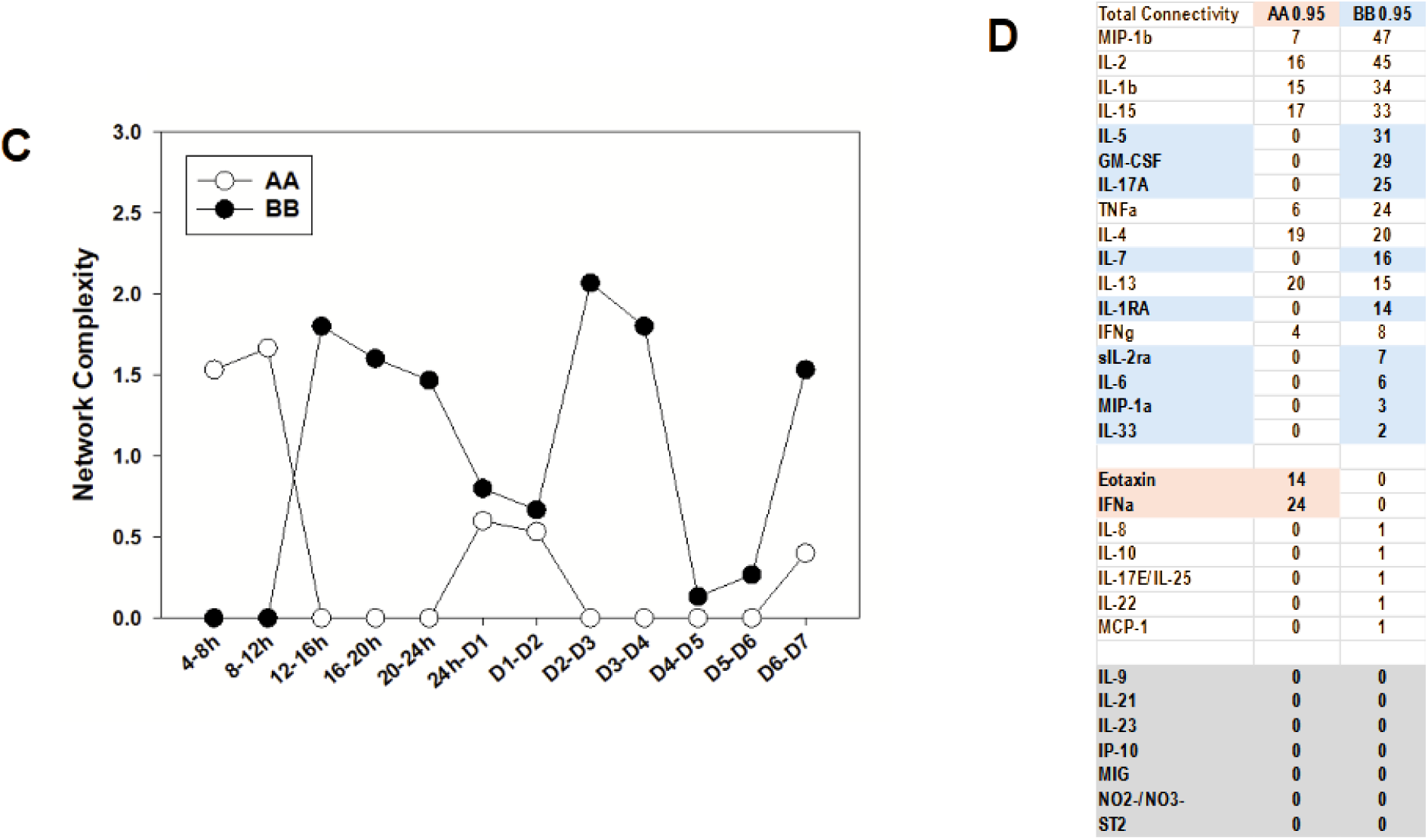
Dynamic network analysis (DyNA) suggests distinct inflammatory programs as a function of homozygous genotype at rs4774381. DyNA results for inflammatory biomolecules from 4h-D7 of rs4774381^AA^ patients **(A)** and rs4774381^BB^ patients **(B)**. A representative DyNA output showing all inflammatory mediators in the networks is also shown to easily identify the mediators. **(C)** Graph of dynamic network complexity over the 4h-D7 time course between rs4774381^AA^ and rs4774381^BB^. **(D)** Table of the total number of connections for each biomolecule for both rs4774381 genotypes.

The overall network connectivity difference between homozygous subgroups was also reflected in the network connectivity of individual inflammatory mediators (Figure 3D). Multiple inflammatory mediators that had zero connections in networks inferred in rs4774381^AA^ patients exhibited high connectivity in rs4774381^BB^ patients; this included IL-17A as well as IL-5, GM-CSF, sIL-2rα, and IL-6 (Figure 3D). In contrast, Eotaxin and IFN-α exhibited the opposite pattern, with zero network connections inferred in rs4774381^BB^ patients and high connectivity in rs4774381^AA^ patients (Figure 3D). Overall, the most connected mediators were MIP-1β and IL-2, both inferred in rs4774381^BB^ patients (Figure 3D).

Since RORA is a key transcription factor associated with Th17 differentiation (38), we next carried out a correlation analysis aimed at inferring Th17 cell subsets as in prior studies (13, 15, 19), noting that identification of Th17 cells in trauma patients using this correlation-based analysis was supported by more direct methods in studies carried out by others (40). This analysis involves correlation of circulating IL-17A with circulating GM-CSF, IL-10, TNF-α, and IL-22 to infer the presence of pathogenic Th17 cells (IL-17A vs. GM-CSF), non-pathogenic Th17 cells (IL-17A vs. IL-10), memory/effector T cells (IL-17A vs. TNF-α), and δγ T cells (IL-17A vs. IL-22), respectively (Figure 4), and suggested distinct patterns of Th17 subpopulations based on statistically significant inter-mediator correlations. Specifically, we found a significant correlation between IL-17A and IL-10 only in rs4774381^AA^ patients (compare Figure 4B and 4F), suggesting the predominance of non-pathogenic (non-inflammatory (38)) Th17 cells in these patients. Both rs4774381^AA^ and rs4774381^BB^ patients exhibited statistically significant correlations between IL-17A and TNF-α (compare Figures 4C and 4G) as well as IL-22 (compare Figures 4D and 4H), suggesting similar levels of memory/effector and δγ T cell responses, respectively. Finally, neither patient subgroup exhibited an upregulation of pathogenic Th17 cells as inferred from non-significant correlations between IL-17A and GM-CSF (compare Figures 4A and 4E). This analysis suggests that upregulation of non-pathogenic Th17 cells is associated with hypo-inflammation following severe injury.

**Figure 4.**
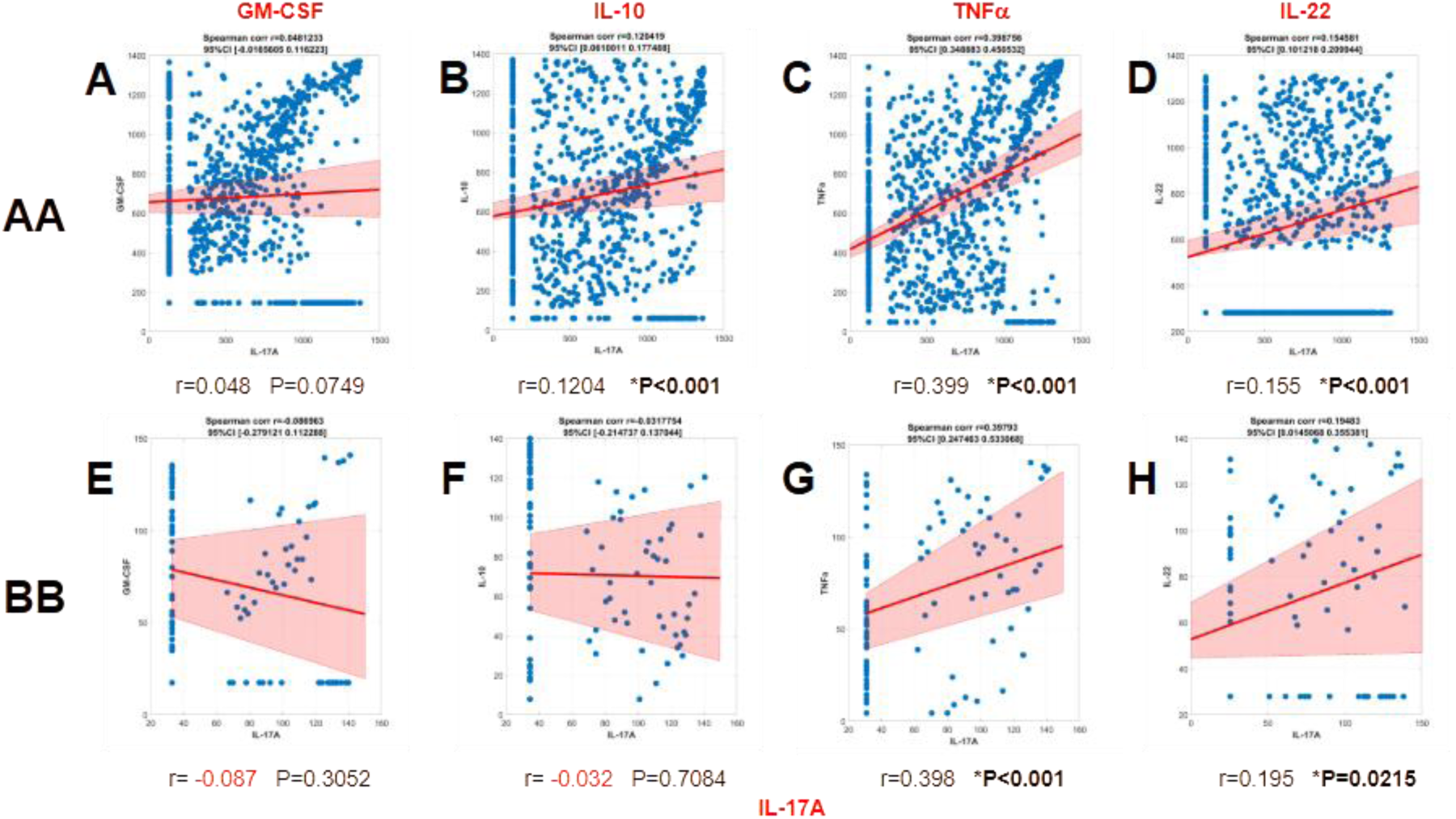
Spearman’s correlations of IL-17A and Th17 differentiation-associated biomolecules: GM-CSF **(A** and **E)**, IL-10 **(B** and **F)**, TNF-α **(C** and **G)** and IL-22 **(D** and **H)**. Panels **(A-D)** are correlations performed for rs4774381^AA^ and panels **(E-H)** are those for rs4774381^BB^.

The SNP rs7918717 in the *RPP38* gene, which codes for a subunit of RNAse P (41) was the second major hub gene identified by FLAME, with four connections. Interestingly, the SNP rs7918717 in the *NMT2* gene was identified as a minor hub gene with one connection also to rs7918717. Patients stratified based on genotype (AA vs. BB) at this SNP exhibited significantly different circulating GM-CSF (p=0.014) as well as MODS trajectories (p=0.024), but not in ICU LOS, total LOS, days on mechanical ventilation, ISS, and AIS (p>0.05, not shown). There was no statistically significant difference in Shock Index between AA and BB (p=0.866).

The SNP rs261537 in the *UBE2D2* gene was the third major hub gene identified by FLAME, with 2 connections. The AA and BB subgroups at this SNP exhibited statistically significant differences in both circulating IFN-γ (p=0.026) and GM-CSF (p=0.042). However, we found no significant differences in any of the clinical characteristics analyzed (not shown).

### 3.2 rs11919443 in the *TM4SF19* gene is associated with distinct trajectories of multiple systemic inflammatory mediators including IL-17A and hypo-inflammation, but not with distinct Th17 cell subsets

We next hypothesized the trauma patient subgroups segregated based on non-hub, rare SNPs associated with genotypically distinct systemic inflammation profiles would also exhibit differences in circulating IL-17A. One such SNP was rs11919443 in the gene coding for transmembrane 4 L six family member 19 (*TM4SF19*). In support of this hypothesis, rs11919443^AA^ patients exhibited significantly elevated levels of circulating IL-17A as compared to rs11919443^BB^ patients (Figure 5A). In addition, the following circulating inflammatory mediators were also significantly different between the two groups by Two-Way ANOVA: IL-6 (p=0.005), IL-7 (p=0.007), IL-8 (p=0.001), IL-10 (p=0.017), MCP-1 (p<0.001) and sST2 (p<0.001). These differences were associated with significantly different clinical outcomes, including longer ICU stay (Figure 5C) and longer time on mechanical ventilation (Figure 5D) in rs11919443^AA^ patients as compared to rs11919443^BB^ patients. In contrast, there was no statistically significant difference in Shock Index between AA and BB (p=0.207),

**Figure 5.**
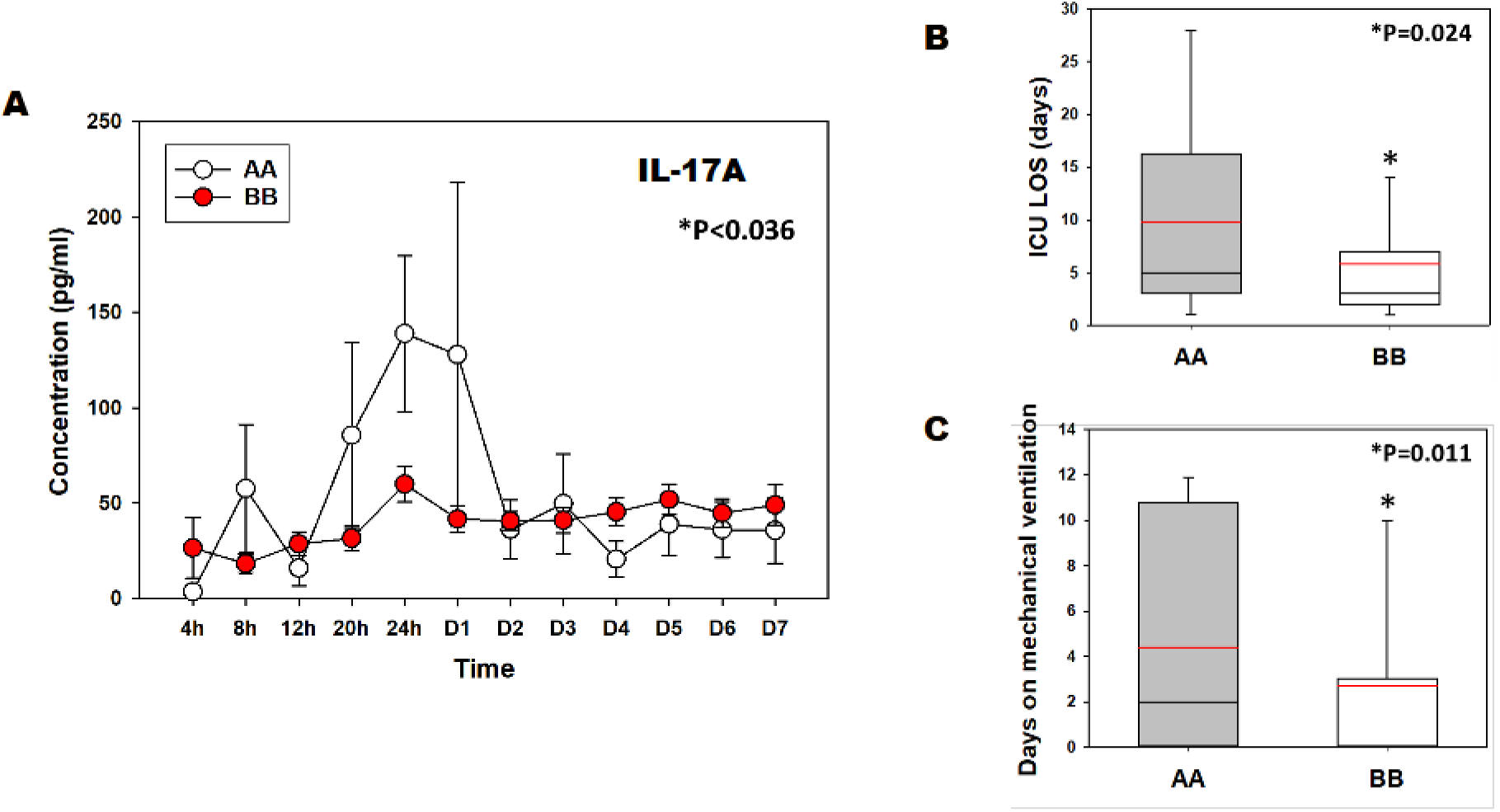
Distinct IL-17A trajectories and clinical outcomes as a function of homozygous genotype at rs11919443. **(A-C)** Post-admission comparison of systemic inflammation and clinical outcomes between rs11919443^AA^ and rs11919443^BB^. **(A)** rs11919443^AA^ patients exhibited a significant spike in IL-17A concentration from 20 hours to 2 days post-injury compared to rs11919443^BB^ patients. **(B-C)** Patients with rs11919443^AA^ had significantly longer ICU stays and required more days on mechanical ventilation than rs11919443^BB^.

Similarly to the analysis of patients stratified based on genotype at rs4774381, inference of dynamic networks in rs11919443^AA^ patients as compared to rs11919443^BB^ patients suggested a striking hypo-inflammatory phenotype, with rs11919443^BB^ patients exhibiting zero inflammatory network connectivity while rs11919443^AA^ patients exhibited fairly robustly connected networks (Figure 6A) (Supplementary Figure 2). The most connected mediators (with 10 connections or more) in rs11919443^AA^ patients were, in descending order, IL-5, IL-7, IL-2, IL-15, IL-1β, GM-CSF, IL-1RA, IL-17A, IFN-γ, and IFN-α (Figure 6B). In agreement with the less important role of IL-17A in these dynamic networks, and in contrast with the analysis carried out on patients stratified based on homozygous genotype rs4774381 at the hub gene *RORA*, rs11919443 patient subgroups exhibited no major inferred differences in their Th17 subsets (Figure 7).

**Figure 6.**
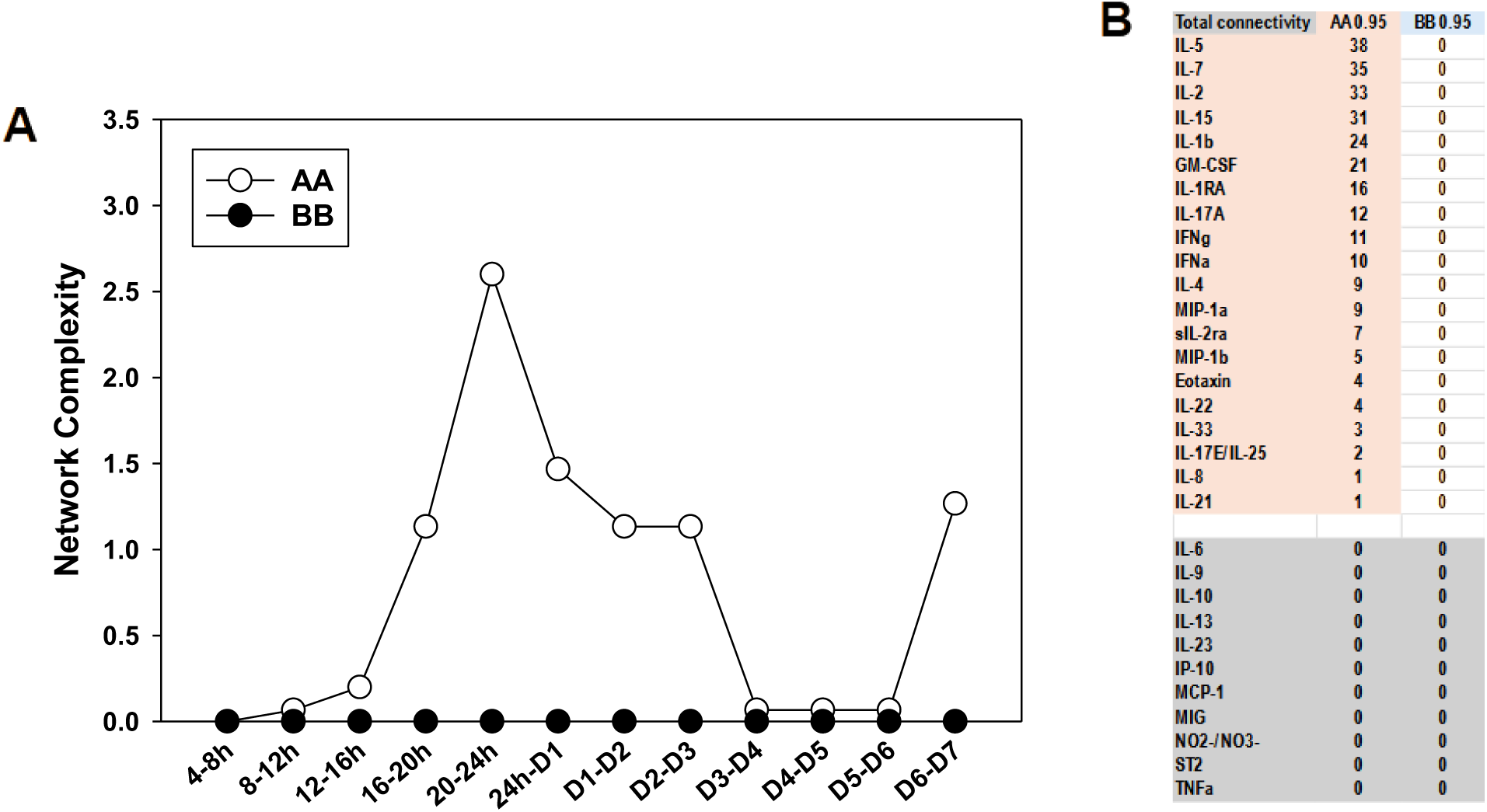
Dynamic network analysis suggests distinct inflammatory programs and the presence of hypo-inflammation as a function of homozygous genotype at rs11919443. **(A)** Graph of dynamic network complexity over the 4h-D7 time course between rs11919443 ^AA^ and rs11919443 ^BB^. **(B)** Table of the total number of connections for each biomolecule for both rs11919443 genotypes.

**Figure 7.**
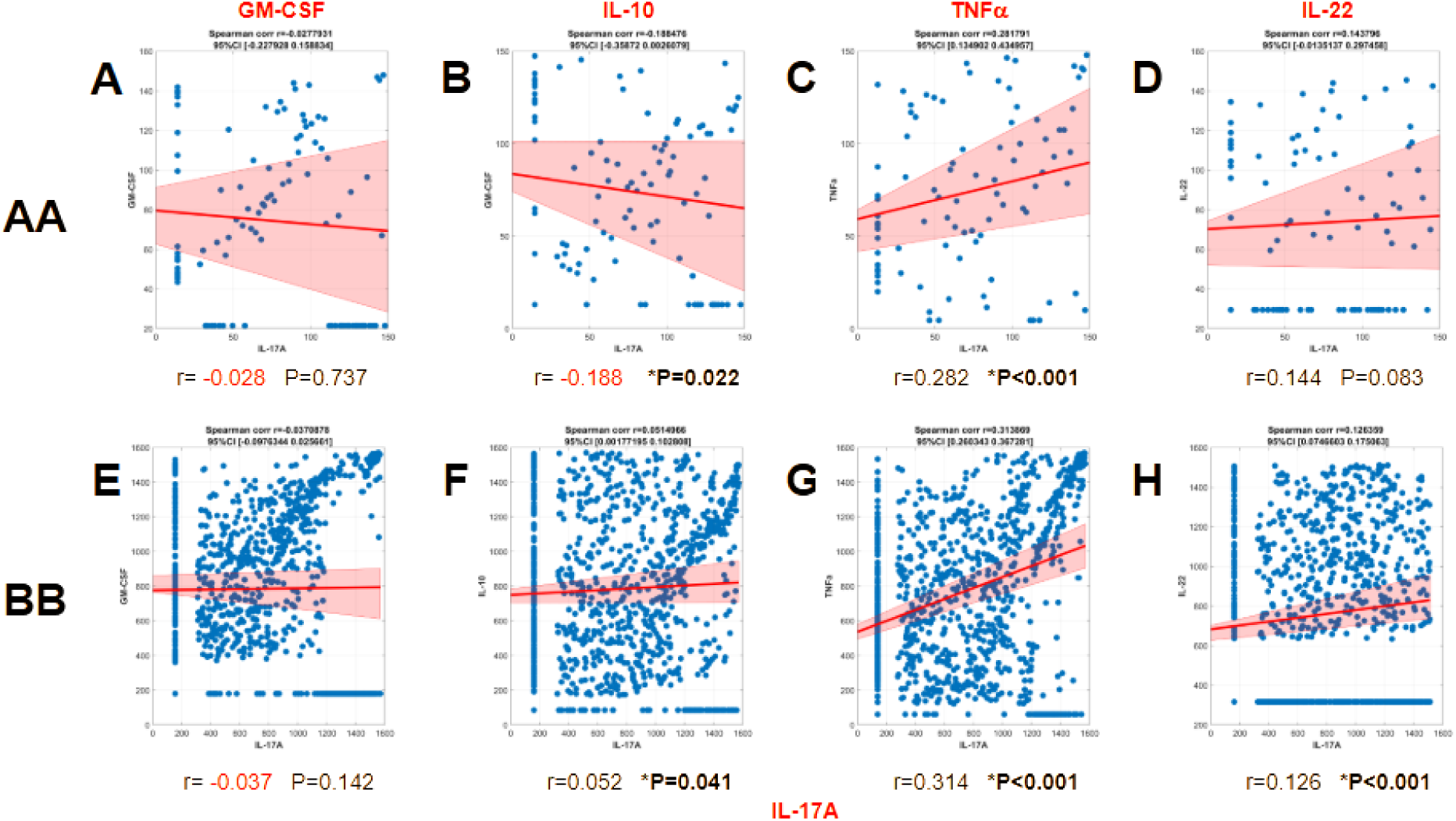
Correlation-based inference of Th17 cell subsets as a function of homozygous genotype at rs11919443. Spearman’s correlations for rs11919443^AA^ **(A-D)** and rs11919443^BB^ **(E-H)** of IL-17A and Th17 differentiation-associated biomolecules: GM-CSF **(A** and **E)**, IL-10 **(B** and **F)**, TNF-α **(C** and **G)**, and IL-22 **(D** and **H)**.

### 3.3 Analysis of SNP networks in a control group defined algorithmically as having genotypically associated differences in clinical outcomes but not circulating inflammatory mediators implicates *SLC2A1*

The 443 rare SNPs with zero statistically significant mediators were entered into FLAME, which returned 261 SNP-gene permutations. When we added the requirement that the control SNPs must have 3-4 statistically different post-traumatic outcomes, the number of rare control SNPs fell to 10 SNPs. Running these SNPs through FLAME resulted in five genes associated with a given SNP within the set, of which four gave results for functional enrichment (Supplementary Table 2). The four genes are *SLC2A1* (associated with rs841852), *IQSEC2* (rs1409117), *ABI3BP* (rs7609779), and *RGS21* (rs7514892). Of these, only *SLC2A1* (solute carrier family 2, facilitated glucose transporter member 1/glucose transporter 1 [GLUT1]), indicated by its SNP rs841852, was a hub gene (Figure 8A-B). Patients segregated based on their genotype at rs841852 (AA vs. BB) did not differ with regard to injury severity (ISS as well as AIS) and Shock Index (p=0.066) but did exhibit statistically significant differences in (Figure 8C-G): ICU length of stay (p< 0.002), total hospital length of stay (p= 0.046), and Marshall MODScore (p< 0.001). Circulating inflammatory mediators in the two genotypes differed significantly (Two-Way ANOVA) in IL-1RA (p= 0.019), IL-7 (p = 0.032), IL-13 (p< 0.001), IL-15 (p= 0.006), MIG (p= 0.024), and sIL-2Ra (p= 0.003) (Figure 9).

**Figure 8.**
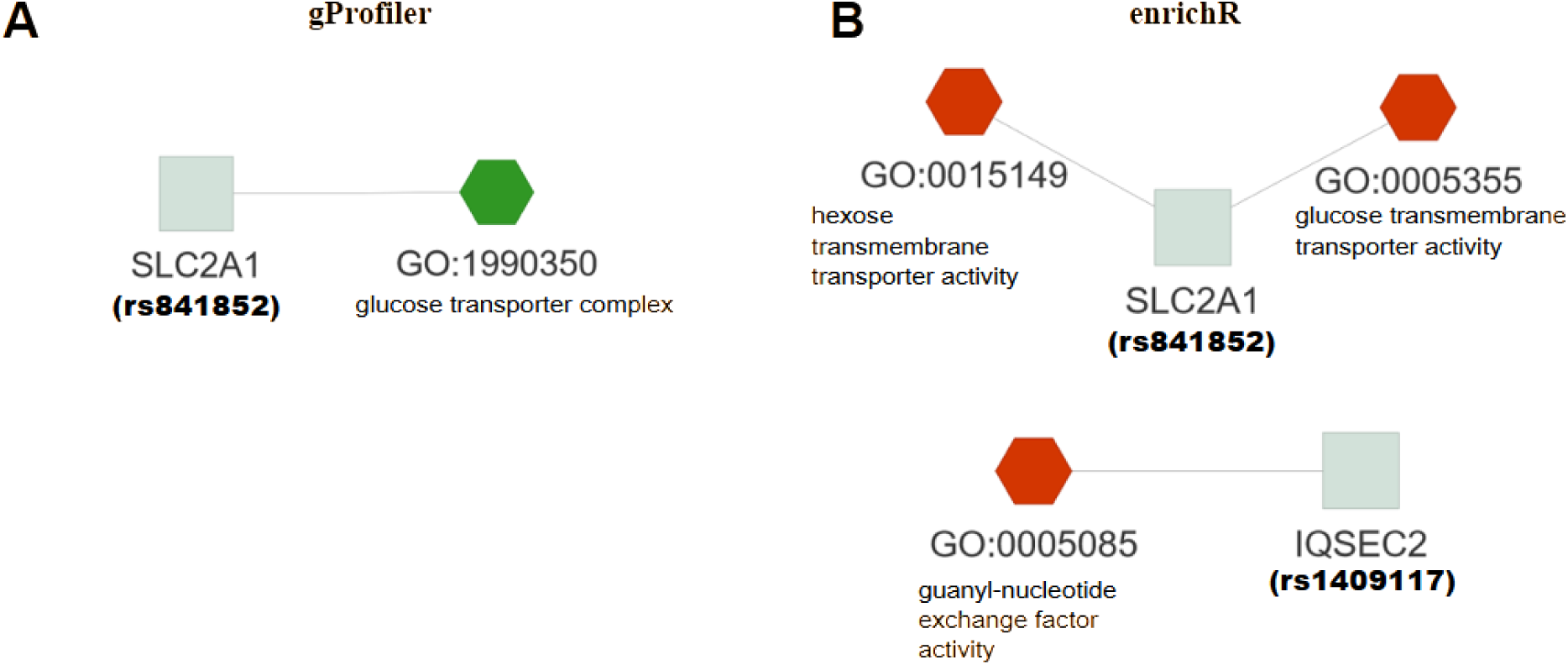
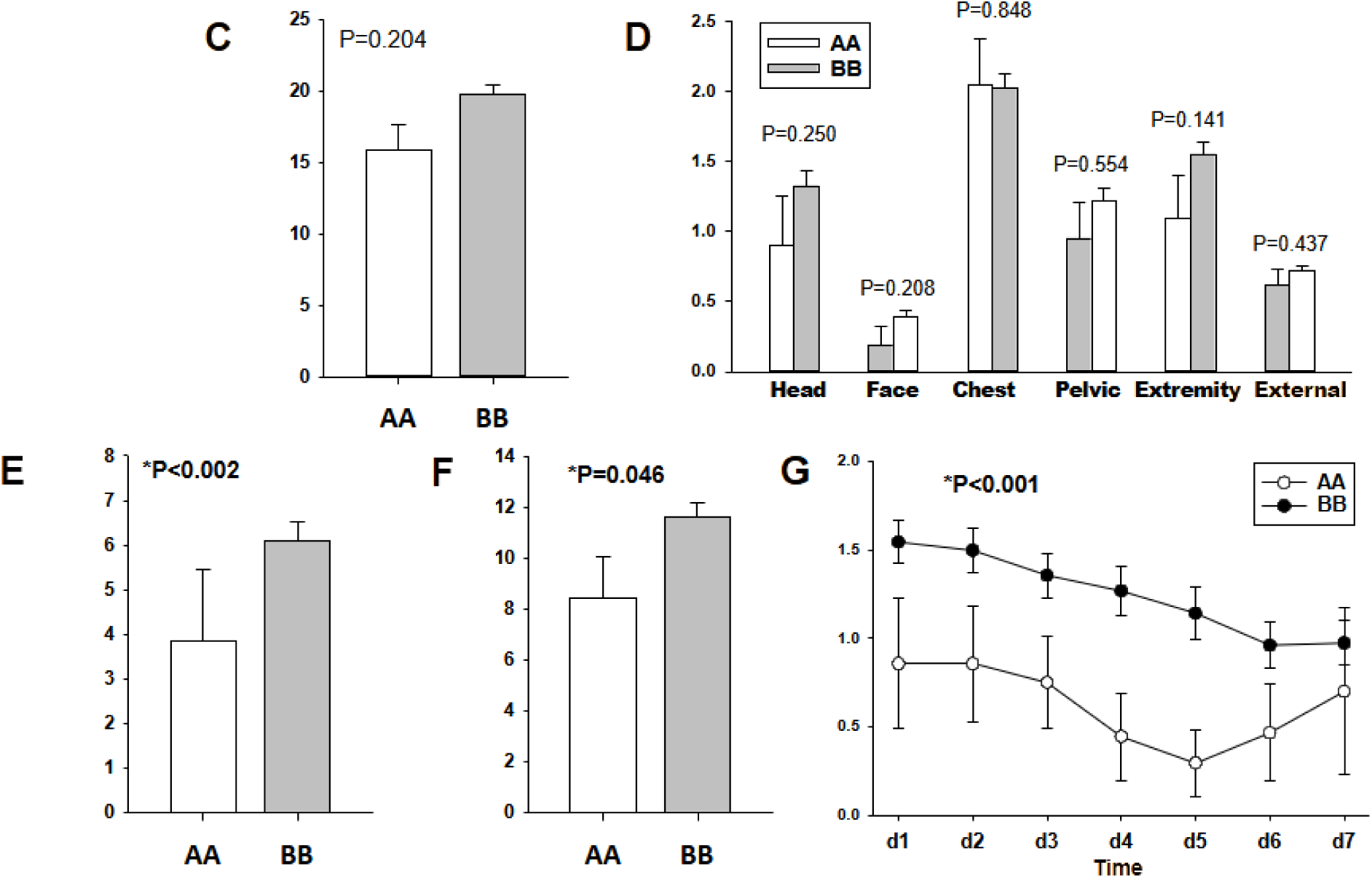
(A-B) FLAME tree plot describing molecular function connectivity of ‘rare’ SNPs with no significant inflammatory biomarker disparity, according to *SNPScanner*. These results were found in the Gene Ontology knowledgebase with the web tools *gProfiler* (A) and *enrichR* (B). Clinical parameter analysis by two-way ANOVA failed to reveal differences in ISS **(C)** and AIS **(D)** between rs841852^AA^ and rs841852^BB^, but implicated significant differences (p < 0.05) for days in the ICU **(E)**, total hospital days **(F)**, and Marshall MODS **(G)**.

**Figure 9.**
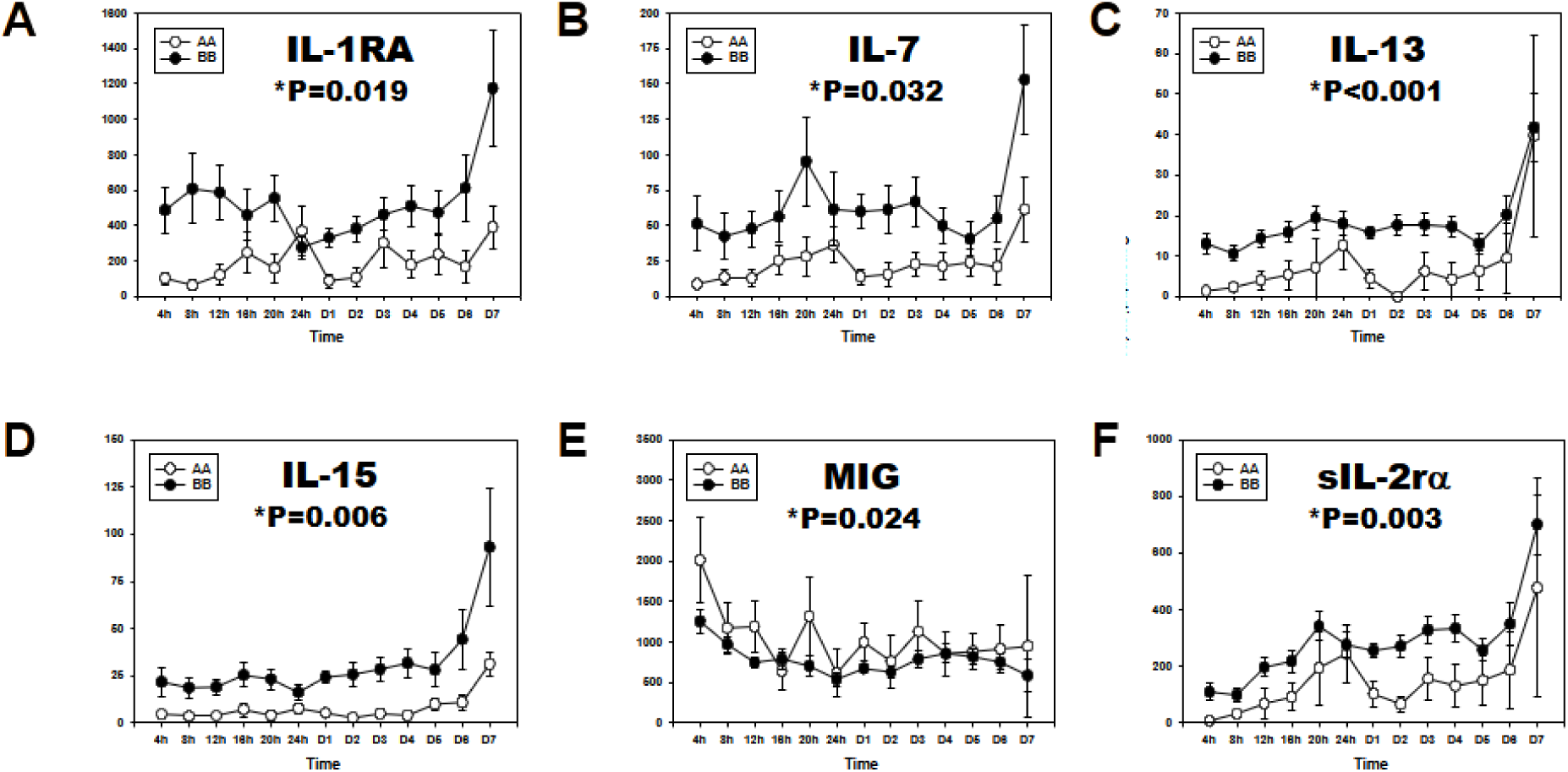
Distinct trajectories of circulating inflammatory mediator mediators as a function of homozygous genotype at rs841852. Statistically significantly different (p < 0.05 by two-way ANOVA) biomarkers: IL-1RA **(A)**, IL-7 **(B)**, IL-13 **(C)**, IL-15 **(D)**, MIG **(E)**, and sIL-2Ra **(F)**.

Similarly to the hypo-inflammation inferred in dynamic networks observed in the other SNP comparison groups, trauma patients segregated based on alternative homozygous genotype at rs841852 exhibited overall low inflammatory network connectivity except in rs841852^AA^ patients at time points from day 4-7 post-hospitalization; rs841852^BB^ patients exhibited zero network connections at all time points (Figure 10A). Mediators with 10 or more connections in rs841852^AA^ patients were, in descending order, MIP-1α, IL-4, IL-5, IFN-γ, IL-7, IL-13, IL-17A, IL-1RA, and IL-33 (Figure 10B; Supplementary Figure 3). Consistent with the hypothesis of hypo-inflammation in rs841852^BB^ patients was a mild inferred upregulation of non-pathogenic Th17 cells while other inferred Th17 subsets were generally similar between patient subgroups (Figure 11).

**Figure 10.**
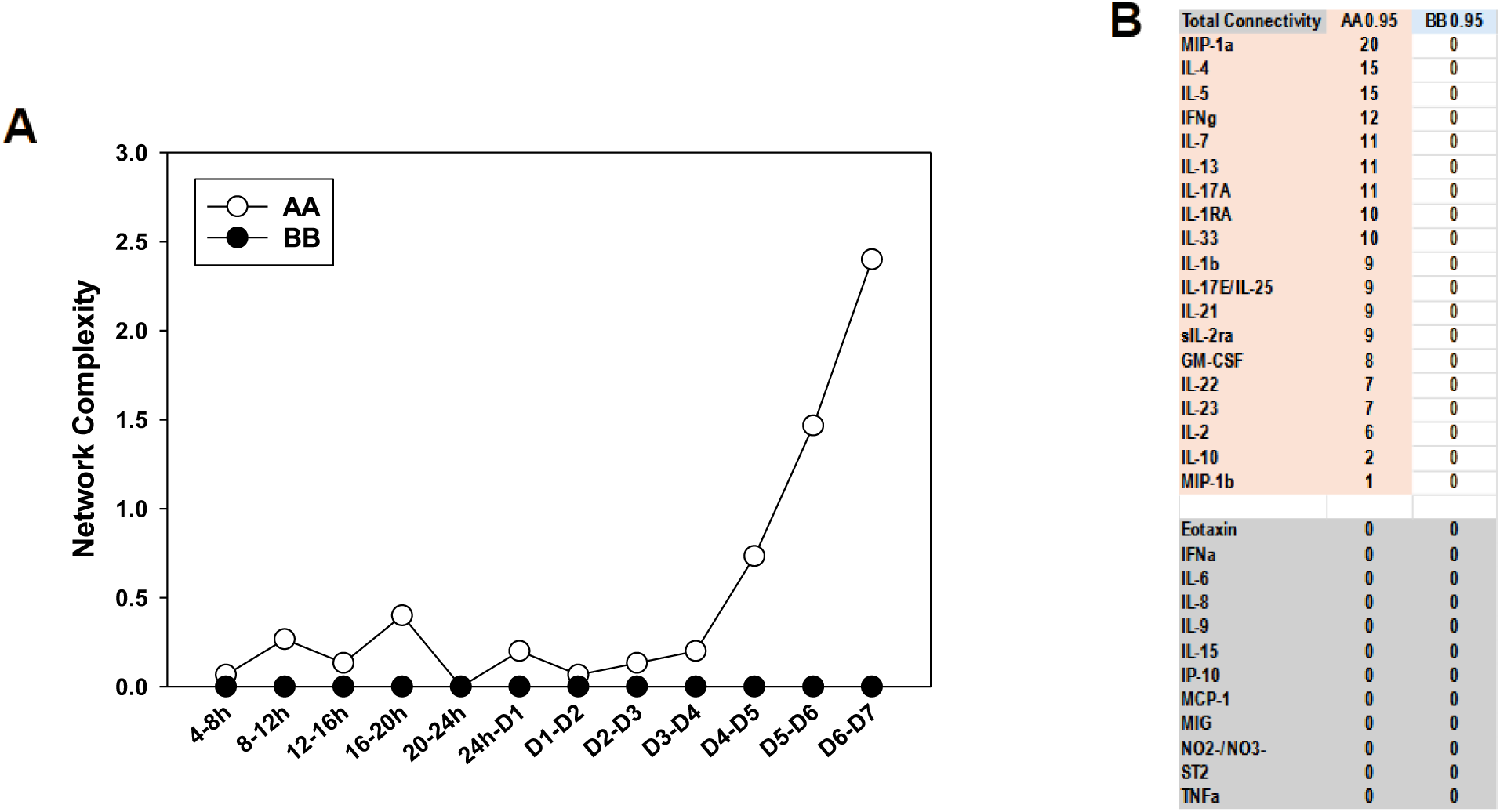
Dynamic network analysis suggests distinct inflammatory programs and the presence of hypo-inflammation as a function of homozygous genotype at rs841852. **(A)** Network complexity of inflammatory biomolecules from 4h-D7 for rs841852 homozygous genotypes. **(B)** Table of total network connections for all inflammatory biomolecules for both genotypes.

**Figure 11.**
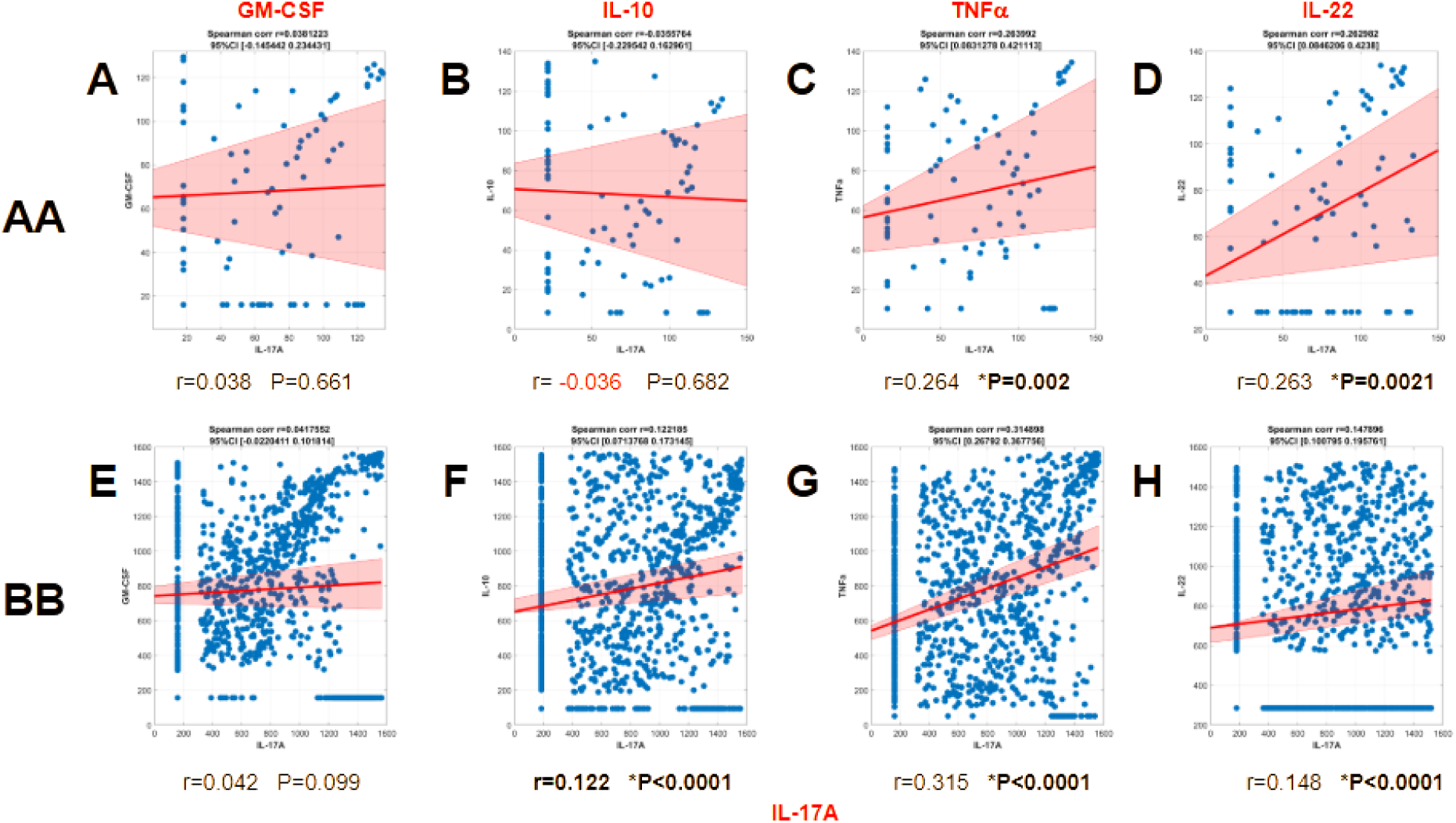
Spearman’s correlations for rs841852 ^AA^ **(A-D)** and rs841852 ^BB^ **(E-H)** of IL-17A and Th17 differentiation-associated biomolecules: GM-CSF **(A** and **E)**, IL-10 **(B** and **F)**, TNF-α **(C** and **G)**, and IL-22 **(D** and **H)**.

## 4. DISCUSSION

The present study extends multiple prior studies linking specific genetic polymorphisms to outcomes of trauma-associated critical illness (19, 22, 23, 25, 42, 43). Specifically, we leveraged novel bioinformatics tools and approaches to implicate the Th17-related RAR-related orphan receptor A (RORA) transcription factor (54) as a major hub gene in networks of SNPs associated with dysregulated systemic inflammation in blunt trauma patients. Segregating blunt trauma survivors based on alternative homozygous genotypes at rs11919443, a non-hub, intronic SNP in the *TM4SF19* gene also identified in this search, yielded subpopulations of patients with distinct trajectories of systemic inflammatory mediators including IL-17A. Furthermore, these subpopulations also differed in key critical illness outcomes but not in core demographics or injury/hemorrhage characteristics, supporting the notion that the differences we identified are due to genetic causes. Our study also suggested *SLC2A1*/*GLUT1* as a hub gene in the responses of trauma patients whose systemic inflammation was not characterized by extensive, genotypically associated differences while still exhibiting genotypically related differences in clinical outcomes. Finally, the present study also highlighted the presence and possible pathophysiologic role of systemic hypo-inflammation, and implicated differential IL-17A production in this process.

Interleukin-17A is a pro-inflammatory cytokine that is produced by Th17 cells, innate lymphoid cells, γδ T cells, and natural killer cells among others (44–50). Interleukin-17A and Th17 cells were studied initially in the context of chronic inflammatory/autoimmune diseases such as rheumatoid arthritis, Crohn’s disease, psoriasis, and systemic lupus erythematosus (SLE) (49, 51). More recently, this pathway has been implicated in acute inflammatory states such as sepsis (52, 53), trauma/hemorrhagic shock (13, 15, 20), COVID-19 (54), wound healing (55), and acute allograft rejection (56).

Interleukin-17A and Th17 cells have also been implicated in the dysregulated inflammation that accompanies adverse outcomes in the context of critical illness, based on initial studies in animal models of sepsis (52, 53) and subsequent studies in human trauma patients (13, 40). Specifically, mortality in trauma-associated critical illness (13) was associated with a predominance of so-called pathogenic Th17 cells (57, 58). In these initial studies documenting an early role for IL-17A in dynamic networks of self-sustaining inflammation, we inferred an upregulation of pathogenic Th17 cells and a concomitant downregulation of nonpathogenic Th17 cells (13). Our initial study associating seven novel SNPs with mortality extended these initial findings and suggested that this genetic predisposition to mortality in the context of trauma-associated critical illness was not determined solely by these SNPs but also required a predominant pathogenic Th17 phenotype that was in turn associated with an overall elevated systemic inflammatory response (19). These results pointed to related, but distinct, roles for genetic predisposition as well as systemic inflammation characterized by distinct Th17 subsets, and this was confirmed in subsequent studies (22). In the present study, we extend these results further with the observation that the flip side of pathogenic Th17-driven, self-sustaining hypo-inflammation is a genetically controlled predisposition to hypo-inflammation characterized by an upregulation of non-pathogenic Th17 cells. We note the early upregulation of systemic IL- 10 and inferred upregulation of non-pathogenic Th17 cells in rs4774381^BB^ patients as compared to rs4774381^AA^ patients. We have suggested a similar phenotype in patients stratified based on a SNP in the *CD55* gene (25). This hypothesis needs to be tested in larger patient cohorts as well as via mechanistic perturbations in appropriate experimental models.

A related hypothesis concerns the IL-10 family cytokine IL-22 (59) and the correlation between IL-17A and IL-22. In this and related studies, we have inferred the presence of γδ T cells based on a positive correlation between IL-17A and IL-22. However, this supposition would need to be tested directly by examining cellular profiles and other markers specific for any of the inferred cell subpopulations, as noted above. It is also possible that the positive correlation between IL-17A and IL-22 reflects an IL-17A-induced upregulation of IL-22 by other cell types. In this regard, recent studies have implicated epithelial-derived IL-22 and related tissue-protective cytokines such as IL-21 and IL-17E/IL-25 in the context of the response to traumatic injury (60). It is intriguing to speculate that IL-17A, derived from any of the cell populations that can produce this cytokine, induces IL-22 from epithelial cells in various tissues and that this process, alone or in concert with upregulation of γδ T cells as well as increases in circulating IL-10, may contribute to systemic hypo-inflammation in some patients. Given the difficulty in teasing apart these processes (especially given the need to assess IL-22 in tissues of human trauma patients), this hypothesis may require studies in animal models or advances in the ability to detect fluxes in cell populations in humans *in situ* (61).

Another intriguing hypothesis that will need to be tested further is that of the apparent “phase shift” in an oscillatory systemic inflammatory response inferred via DyNA when stratifying trauma patients based on homozygous genotype at rs4774381. In unpublished studies, we have inferred similar dynamic network oscillations when examining the systemic inflammatory responses of spinal cord injury patients segregated based on the location of their spinal cord injury (T6 and above vs. below T6; R. Zamora, E. Robbins, W. Huang, Y. Vodovotz, and G. Sowa, unpublished), which raises the possibility of neural involvement in this phenomenon. In support of this hypothesis, we have previously documented a role for the chemokine IP-10/CXCL10 as a driver of IL-10 and systemic hypo-inflammation when comparing trauma patients with vs. without spinal cord injury (62).

rs11919443 in the *TM4SF19* gene was another key SNP inferred to be related to dysregulated systemic inflammation (including distinct IL-17A time courses as a function of genotype at this SNP). Specifically, patients with the homozygous AA genotype of rs11919443 exhibited significantly longer ICU length of stay and requirement for mechanical ventilation (along with a trend towards longer total hospital length of stay) as well as higher circulating IL-17A levels as compared to patients with the alternative homozygous (BB) genotype. Transmembrane 4 L six family member 19 (TM4SF19) is a lysosomal protein that suppresses acidification; its inactivation was found to block the increase in accumulation of macrophages in a murine adipose tissue and thus was associated with reduced inflammation (63). *In vitro* studies suggested that TM4SF19 exacerbates the effects of bacterial lipopolysaccharide on human venous endothelial cells by impacting the expression of cell junction proteins (64). Thus, TM4SF19 appears to exert pro-inflammatory effects, albeit indirectly.

One such indirect mechanism appears to involve the chemokine CXCL2, based on a recent bioinformatics study linking TM4SF19 and CXCL2 in the context of childhood obesity (65). Interleukin-17A induced CXCL2 among its many actions (66), and bioinformatics analyses have therefore placed CXCL2 in the IL-17A signaling pathway (67, 68). Furthermore, IL-17A synergizes with TNF-α to drive endothelial expression of CXCL2 and subsequent neutrophil recruitment (69).

A key aspect of the present study is the focus on a control group of SNPs selected based on the following criteria: 1) absence of statistically significant differences in circulating inflammatory mediators between subgroups of trauma patients stratified on the basis of their homozygous genotype at a given SNP, and 2) presence of statistically significant differences in clinical outcomes in these same patients. Once SNPs that met these criteria were selected, FLAME analysis was used to define hub genes. This analysis identified *SLC2A1* as the main hub gene. SLC2A1 (solute carrier family 2, facilitated glucose transporter member 1/glucose transporter 1 [GLUT1]) is the main glucose transporter (70), in which there has been extensive interest due to the emerging role of glucose metabolism in autoimmunity (71, 72), cancer (73, 74), as well as sepsis-induced critical illness (75). Our findings support a role for differential glucose transport in organ dysfunction in the context of trauma-induced critical illness as well.

Our study has several limitations. These include the need to assess more SNPs and other genetic variants in larger, more diverse patient cohorts; to measure more mediators to better define the impact of the specific SNPs assessed herein on the immuno-inflammatory response; and to assess other ‘omics layers to better delineate the host’s pathophysiological response in the context of genetic predisposition. Computational modeling – adding mechanistic modeling to the present study’s data-driven modeling approaches (61) – would also serve an important role in any attempt to connect our findings to the host’s pathophysiology following severe traumatic injury and would also help address the network of interaction among SNPs (76, 77). Finally, our conclusions are based on associations rather than direct mechanistic work, though our initial study documenting the role of IL-17A in the injury response included supportive mechanistic studies involving neutralization of IL-17A in a mouse model of trauma/hemorrhage (13).

Despite these limitations, the present study suggests a clear role for nuanced Th17 response in the context of trauma-associated systemic inflammation and critical illness. Multiple cell types can express IL-17A, including Th17 cells, δγ T cells, and innate lymphoid cells (ILCs) (44–50). In the context of trauma-associated critical illness or experimental models of trauma/hemorrhage, IL-17A production is associated with morbidity and mortality (13, 15, 19–21). While studies have implicated both Th17 cells (13, 40) and ILCs (78) in this pathophysiology, the present study points to RORA and Th17 differentiation as a key genetically regulated aspect of this response; future studies will hopefully address the relative contributions of these distinct cell populations.

## Data Availability

Patient data used in the present study are confidential and under Health Insurance Portability and Accountability Act protection. All analysis data produced in the present study are available upon reasonable request to the authors. *SNPScanner* code is available online at https://github.com/fayteneldehaibi/SNPScanner.

## ABBREVIATIONS

ABI3BP: ABI family member 3 binding protein
AIS: Abbreviated Injury Scale
ANOVA: Analysis of Variance
DyNA: Dynamic Network Analysis
GLUT1: glucose transporter 1
GM-CSF: granulocyte-macrophage colony stimulating factor
GO: Gene Ontology Knowledgebase
ICU: intensive care unit
IFN: interferon
IL: Interleukin
ILC: innate lymphoid cell
IP-10: IFN-γ inducible protein (CXCL10)
IL: interleukin
IQSEC2: IQ motif and Sec7 domain ArfGEF 2
ISS: Injury Severity Score
KEGG: Kyoto Encyclopedia of Genes and Genomics
LOS: length of stay
LYPD4: LY6/PLAUR domain containing 4
MCP: monocyte chemotactic protein
MIG: monokine induced by gamma interferon (CXCL9)
MIP: macrophage inflammatory protein
MODS: Multiple Organ Dysfunction Syndrome
NMT2: N-myristoyltransferase 2
RGS21: regulator of G protein signaling 21
RPP38: ribonuclease P/MRP subunit p38
RORA: RAR-related orphan receptor A
SLC2A1: solute carrier family 2, facilitated glucose transporter member 1
SNP: single nucleotide polymorphism
sST2: soluble suppression of tumorigenicity-2
TM4SF19: transmembrane 4 L six family member 19
TNF-α: tumor necrosis factor α
UBE2D2: ubiquitin conjugating enzyme E2 D2

## FUNDING

This work was supported by Department of Defense grant W81XWH-18-2-0051 and Defense Advanced Research Projects Agency grant D20AC00002.

## ACKNOWLEDGMENTS

This work was made possible by the Department of Defense and the Defense Advanced Research Projects Agency. The manuscript was first published as a preprint to medRxiv.org.

## AUTHOR CONTRIBUTIONS

FE: Writing, software development, data analysis, investigation, visualization. RZ: Writing, data analysis, visualization. JY: Inflammatory mediator arrays, genomic analysis. TB: Curation of patient data, review and editing. YZ: Conceptualization, writing, review and editing, project administration, funding acquisition. This manuscript has been read and approved by all authors.

## CONFLICT OF INTEREST

The authors declare that the research was conducted in the absence of any commercial or financial relationships that could be construed as a potential conflict of interest.

## Notes

### Competing Interest Statement

The authors have declared no competing interest.

### Author Declarations

The Institutional Review Board (Protocol No. MOD08010232-19 / PRO08010232) of the University of Pittsburgh gave ethical approval for use of patient data in this work.

### Summary of Updates

We have given the tables subsection headers (ie. A, B, etc.) to allow the tables to still be large yet organized.

