## Supplementary Figures for "Network Analysis of Rare Single-Nucleotide Polymorphisms Suggests a Central Role for Type 17 Immune Responses in Trauma-induced, Genotypically Associated Hypo-Inflammation and Critical Illness"

**Supplementary Figure 1.** Enlarged view of DyNA graph to clearly show the labels of the inflammatory biomolecules analyzed. The arrangement of these biomolecules is preserved in all DyNA figures with this paper. Starting at the top and moving clockwise, the biomolecules are: Eotaxin, GM-CSF, IFN-α, IFN-γ, IL-1β, IL-1RA, IL-2, IL-4, IL-5, IL-6, IL-7, IL-8, IL-9, IL-10, IL-13, IL-15, IL-17A, IL-17E/IL-25, IL-21, IL-22, IL-23, IL-33, IP-10, MCP-1, MIG, MIP-1α, MIP-1β, $NO_2^-/NO_3^-$, sIL-2Rα, ST2, and TNF-α.

**Supplementary Figure 2.** DyNA of inflammatory biomolecule connections for $rs11919443^{AA}$ (A) and $rs11919443^{BB}$ (B). No connections were observed between any biomolecules at time for $rs11919443^{BB}$. A representative DyNA output showing all inflammatory mediators in the networks is also shown to easily identify the mediators.

**Supplementary Figure 3.** DyNA of inflammatory biomolecules for $rs841852^{AA}$ (A) and $rs841852^{BB}$. A representative DyNA output showing all inflammatory mediators in the networks is also shown to easily identify the mediators.

Suppl. Figure 1

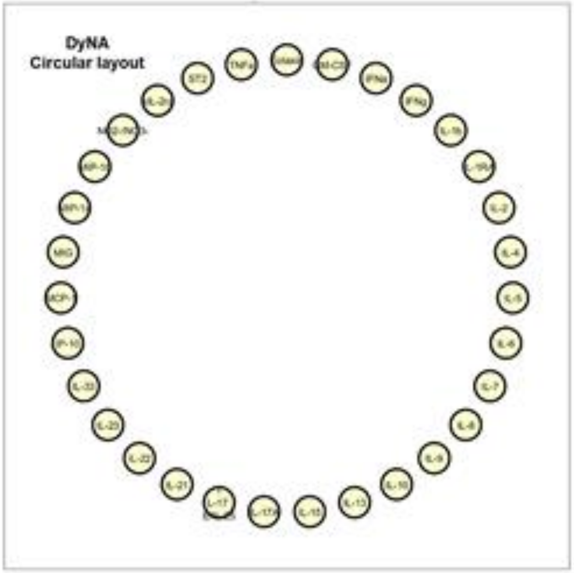

Suppl. Fig. 2

**A**

**AA**

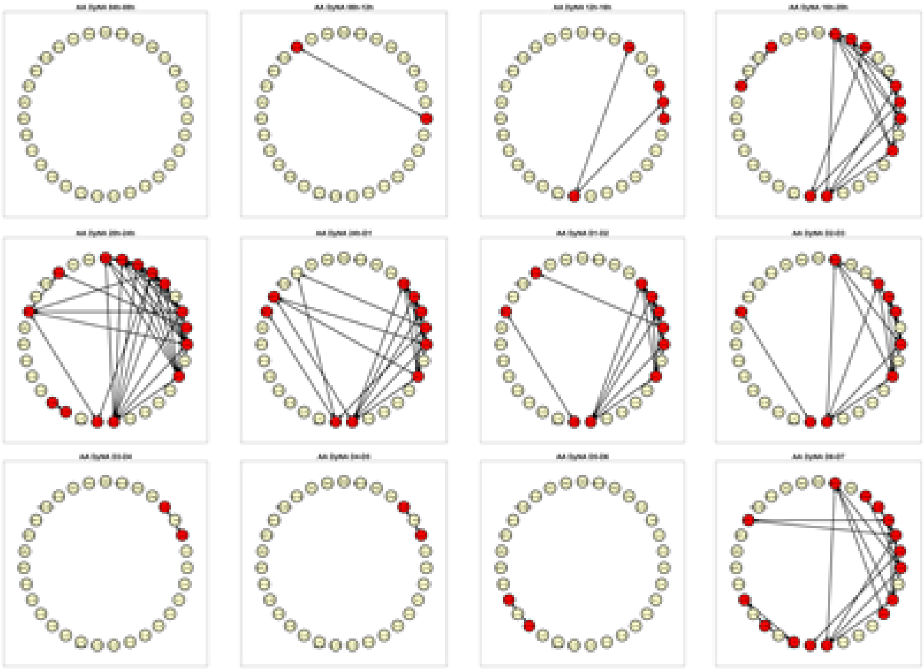

Suppl. Fig. 2

B

BB

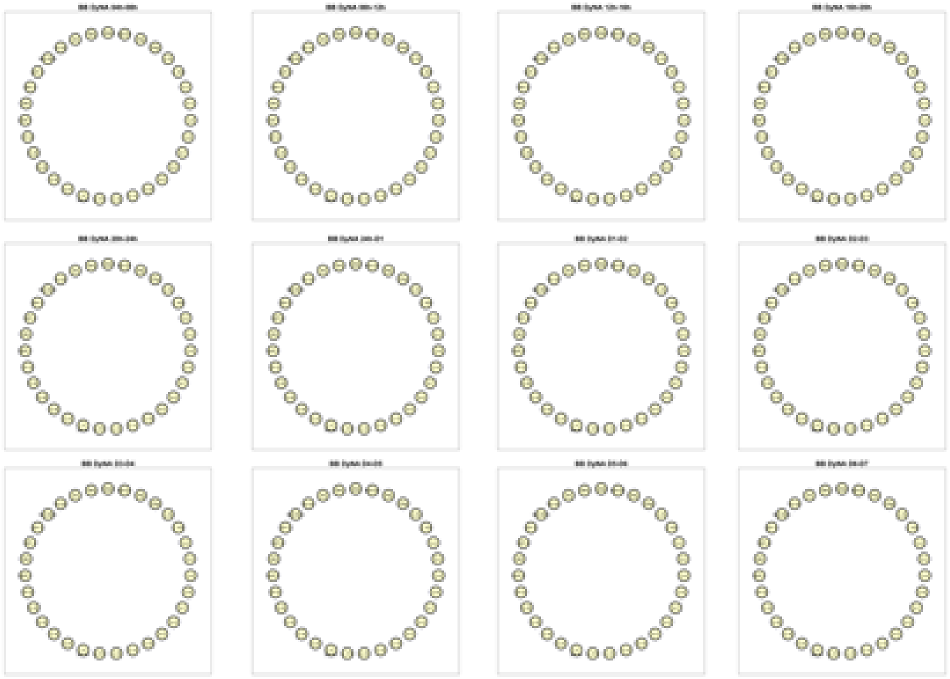

Suppl. Fig. 3

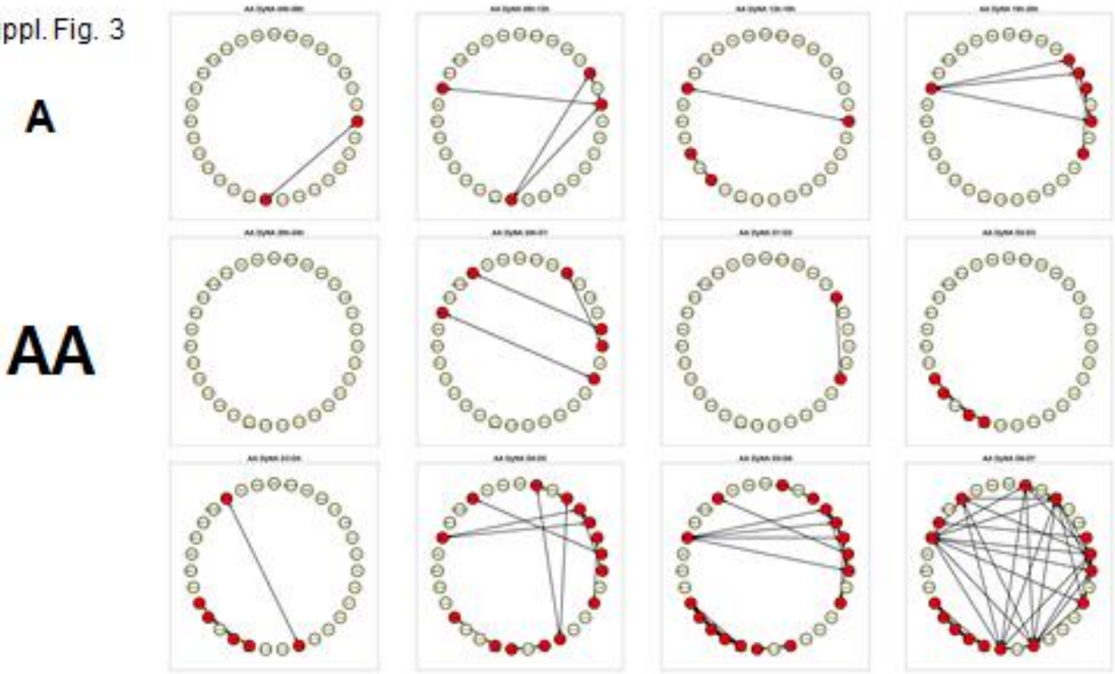

Suppl. Fig. 3

**B**

**BB**

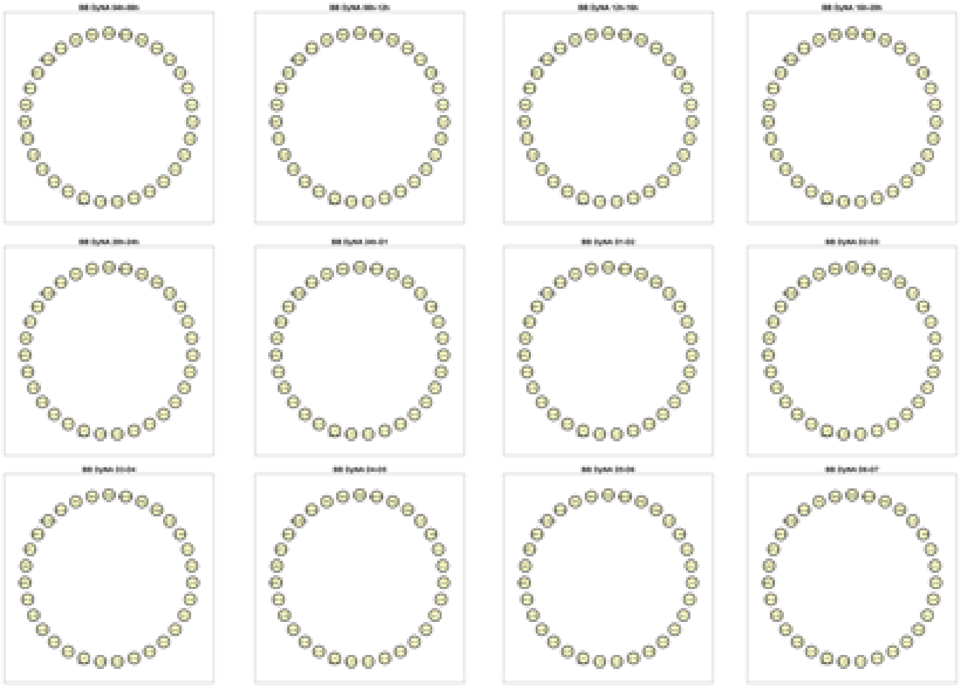
