## Supplementary Tables for "Network Analysis of Rare Single-Nucleotide Polymorphisms Suggests a Central Role for Type 17 Immune Responses in Trauma-induced, Genotypically Associated Hypo-Inflammation and Critical Illness"

Supplementary Table 1. Evenly distributed control SNPs with 3-4 statistically distinct post-traumatic outcomes between AA and BB genotypes ($p < 0.05$).

| SNP | ICU Days | Total Hospital Days | Vent Days | Marshall Mods | Gene Found with FLAME |
|---|---|---|---|---|---|
| rs992708 | ✓ | ✓ | ✓ | ✓ | SNX7 |
| rs10865443 | ✓ | ✓ | ✓ | ✓ | CTNNA2 |
| exm2263860 | ✓ | ✓ | ✓ | ✓ | --- |
| rs13078747 | ✓ | ✓ | ✓ | ✓ | CACNA1D |
| rs704716 | ✓ | ✓ | ✓ | ✓ | B3GALNT2 |
| rs4771918 | ✓ | | ✓ | ✓ | CLDN10, CLDN10-AS1 |
| rs6932393 | ✓ | ✓ | | ✓ | CDKAL1 |
| exm-rs89107 | | ✓ | ✓ | ✓ | --- |
| rs12986520 | | ✓ | ✓ | ✓ | LINC00276 |
| rs1671471 | ✓ | ✓ | | ✓ | --- |
| rs896973 | ✓ | | ✓ | ✓ | TPCN2, ENSG00000287725 |
| rs4685385 | ✓ | ✓ | | ✓ | --- |
| rs7701186 | ✓ | ✓ | | ✓ | --- |
| rs7967371 | ✓ | ✓ | | ✓ | --- |
| rs719680 | ✓ | | ✓ | ✓ | --- |
| rs12127469 | | ✓ | ✓ | ✓ | ZRANB2-DT |

Supplementary Table 2. Rare control SNPs with 3-4 statistically distinct post-traumatic outcomes between AA and BB genotypes ($p < 0.05$).

| SNP | ICU Days | Total Hospital Days | Vent Days | Marshall Mods | Gene Found with FLAME |
|---|---|---|---|---|---|
| rs1409117 | ✓ | ✓ | ✓ | ✓ | IQSEC2 |
| rs2388252 | ✓ | ✓ | ✓ | ✓ | --- |
| rs12450000 | ✓ | ✓ | | ✓ | --- |
| rs4144013 | | ✓ | ✓ | ✓ | PPRG1 |
| rs6609469 | ✓ | ✓ | | ✓ | --- |
| rs9558839 | ✓ | ✓ | | ✓ | --- |
| rs7609779 | ✓ | | ✓ | ✓ | ABI3BP |
| rs767478 | ✓ | ✓ | | ✓ | --- |
| rs841852 | ✓ | ✓ | | ✓ | SLC2A1 |
| rs7514892 | | ✓ | ✓ | ✓ | RGS21 |

Supplementary Table 3. FLAME results of evenly distributed control SNPs with *gProfiler*.

| Source | Term_ID | Function | P-value | Enrichment Score % | Positive Hits |
|---|---|---|---|---|---|
| GO:MF | GO:0022832 | voltage-gated channel activity | 3.28E-03 | 1.61 | CACNA1D, ENSG00000287725, TPCN2 |
| GO:MF | GO:0005245 | voltage-gated calcium channel activity | 5.22E-03 | 3.77 | CACNA1D, TPCN2 |
| GO:MF | GO:0022836 | gated channel activity | 5.22E-03 | 0.95 | CACNA1D, ENSG00000287725, TPCN2 |
| GO:MF | GO:0086059 | voltage-gated calcium channel activity involved SA node cell action potential | 5.78E-03 | 50 | CACNA1D |
| GO:MF | GO:0022803 | passive transmembrane transporter activity | 5.78E-03 | 0.59 | CACNA1D, ENSG00000287725, TPCN2 |
| GO:MF | GO:0005216 | monoatomic ion channel activity | 5.78E-03 | 0.68 | CACNA1D, ENSG00000287725, TPCN2 |
| GO:MF | GO:0015267 | channel activity | 5.78E-03 | 0.59 | CACNA1D, ENSG00000287725, TPCN2 |
| GO:MF | GO:0035598 | N6-threonylcarbomyladenosine methylthiotransferase activity | 5.78E-03 | 100 | CDKAL1 |
| GO:MF | GO:0050497 | alkylthioltransferase activity | 5.78E-03 | 50 | CDKAL1 |
| GO:MF | GO:0035596 | methylthiotransferase activity | 5.78E-03 | 50 | CDKAL1 |
| GO:MF | GO:0061712 | tRNA (N(6)-L-threonylcarbamoyladenosine(37)-C(2))-methylthiotransferase | 5.78E-03 | 100 | CDKAL1 |
| GO:MF | GO:0005262 | calcium channel activity | 7.04E-03 | 1.57 | CACNA1D, TPCN2 |
| GO:MF | GO:0097682 | intracellular phosphatidylinositol-3,5-bisphosphate-sensitive monatomic cation channel activity | 7.21E-03 | 33.33 | TPCN2 |
| GO:MF | GO:0015085 | calcium ion transmembrane transporter activity | 7.85E-03 | 1.38 | CACNA1D, TPCN2 |
| GO:MF | GO:0022843 | voltage-gated monoatomic cation channel activity | 8.47E-03 | 1.28 | CACNA1D, TPCN2 |
| GO:MF | GO:0086007 | voltage-gated calcium channel activity involved in cardiac muscle cell action potential | 9.19E-03 | 20 | CACNA1D |
| GO:MF | GO:0072345 | NAADP-sensitive calcium-release channel activity | 9.19E-03 | 20 | TPCN2 |
| GO:MF | GO:0015075 | monoatomic ion transmembrane transporter activity | 9.42E-03 | 0.41 | CACNA1D, ENSG00000287725, TPCN2 |
| GO:MF | GO:0005244 | voltage-gated monoatomic ion channel activity | 9.42E-03 | 1.09 | CACNA1D, TPCN2 |

| | | | | | |
|---|---|---|---|---|---|
| GO:MF | GO:0035091 | phosphatidylinositol binding | 1.96E-02 | 0.72 | SNX7, TPCN2 |
| GO:MF | GO:0022857 | transmembrane transporter activity | 2.48E-02 | 0.27 | CACNA1D, ENSG00000287725, TPCN2 |
| GO:MF | GO:0005261 | monoatomic cation channel activity | 2.47E-02 | 0.6 | CACNA1D, TPCN2 |
| GO:MF | GO:0030506 | ankyrin binding | 2.47E-02 | 5.26 | CACNA1D |
| GO:MF | GO:0015278 | calcium-release channel activity | 2.47E-02 | 5.56 | TPCN2 |
| GO:MF | GO:0015280 | ligand-gated sodium channel activity | 2.47E-02 | 5.88 | TPCN2 |
| GO:MF | GO:0005215 | transporter activity | 2.90E-02 | 0.24 | CACNA1D, ENSG00000287725, TPCN2 |
| GO:MF | GO:0051393 | alpha-actinin binding | 2.90E-02 | 3.57 | CACNA1D |
| GO:MF | GO:0008331 | high voltage-gated calcium channel activity | 2.87E-02 | 4.17 | CACNA1D |
| GO:MF | GO:0080025 | phosphatidylinositol-3,5-bisphosphate binding | 2.90E-02 | 3.7 | TPCN2 |
| GO:MF | GO:0099604 | ligand-gated calcium channel activity | 2.90E-02 | 3.7 | TPCN2 |
| GO:MF | GO:0005217 | intracellular ligand-gated monoatomic ion channel activity | 3.01E-02 | 3.33 | TPCN2 |
| GO:MF | GO:0046873 | metal ion transmembrane transporter activity | 3.06E-02 | 0.45 | CACNA1D, TPCN2 |
| GO:MF | GO:0008376 | acetylgalactosaminyltransferase activity | 3.20E-02 | 2.94 | B3GALNT2 |
| GO:MF | GO:0042805 | actinin binding | 3.28E-02 | 2.7 | CACNA1D |
| GO:MF | GO:0005543 | phospholipid binding | 3.28E-02 | 0.41 | SNX7, TPCN2 |
| GO:MF | GO:0005272 | sodium channel activity | 3.71E-02 | 2.33 | TPCN2 |
| GO:MF | GO:0051539 | 4 iron, 4 sulfur cluster binding | 3.77E-02 | 2.22 | CDKAL1 |
| GO:MF | GO:0022890 | inorganic cation transmembrane transporter activity | 4.11E-02 | 0.35 | CACNA1D, TPCN2 |
| KEGG | map05412 | Arrhythmogenic right ventricular cardiomyopathy | 4.27E-02 | 2.38 | CACNA1D, CTNNA2 |
| KEGG | map04670 | Leukocyte transendothelial migration | 4.27E-02 | 1.75 | CLDN10, CTNNA2 |
| GO:MF | GO:0008324 | monoatomic cation transmembrane transporter activity | 4.54E-02 | 0.33 | CACNA1D, TPCN2 |

Supplementary Table 4. FLAME results of evenly distributed control SNPs with *enrichR*.

| Source | Term_ID | Function | P-value | Enrichment Score % | Positive Hits |
|---|---|---|---|---|---|
| GO:MF | GO:0005245 | voltage-gated calcium channel activity | 2.14E-03 | 5.41 | CACNA1D, TPCN2 |
| GO:MF | GO:0005262 | calcium channel activity | 4.92E-03 | 2.38 | CACNA1D, TPCN2 |

| | | | | | |
|---|---|---|---|---|---|
| GO:MF | GO:0022843 | voltage-gated cation channel activity | 4.92E-03 | 2.06 | CACNA1D, TPCN2 |
| GO:MF | GO:0086007 | voltage-gated calcium channel activity involved in cardiac muscle cell action potential | 8.09E-03 | 20 | CACNA1D |
| GO:MF | GO:0072345 | NAADP-sensitive calcium-release channel activity | 8.09E-03 | 20 | TPCN2 |
| GO:MF | GO:0008331 | high voltage-gated calcium channel activity | 1.35E-02 | 10 | CACNA1D |
| KEGG | map05412 | Arrhythmogenic right ventricular cardiomyopathy (ARVC) | 1.72E-02 | 2.7 | CACNA1D, CTNNA2 |
| GO:MF | GO:0030506 | ankyrin binding | 1.88E-02 | 5.26 | CACNA1D |
| GO:MF | GO:0042805 | actinin binding | 1.88E-02 | 4.76 | CACNA1D |
| GO:MF | GO:0015278 | calcium-release channel activity | 1.88E-02 | 5.88 | TPCN2 |
| GO:MF | GO:0051393 | alpha-actinin binding | 1.94E-02 | 4.17 | CACNA1D |
| GO:MF | GO:0008376 | acetylgalactosaminyltransferase activity | 1.98E-02 | 3.7 | B3GALNT2 |
| KEGG | map04670 | Leukocyte transendothelial migration | 2.01E-02 | 1.69 | CLDN10, CTNNA2 |
| KEGG | map04530 | Tight junction | 2.01E-02 | 1.44 | CLDN10, CTNNA2 |
| GO:BP | GO:0086015 | SA node cell action potential | 3.22E-02 | 14.29 | CACNA1D |
| GO:BP | GO:0086046 | membrane depolarization during SA node cell action potential | 3.22E-02 | 20 | CACNA1D |
| GO:BP | GO:0045762 | positive regulation of adenylate cyclase activity | 3.22E-02 | 14.29 | CACNA1D |
| GO:BP | GO:0060372 | regulation of atrial cardiac muscle cell membrane repolarization | 3.22E-02 | 14.29 | CACNA1D |
| GO:BP | GO:0060134 | prepulse inhibition | 3.22E-02 | 16.67 | CTNNA2 |
| GO:BP | GO:0034316 | negative regulation of Arp2/3 complex-mediated actin nucleation | 3.22E-02 | 16.67 | CTNNA2 |
| GO:BP | GO:0051823 | regulation of synapse structural plasticity | 3.22E-02 | 20 | CTNNA2 |
| GO:BP | GO:0019065 | receptor-mediated endocytosis of virus by host cell | 3.22E-02 | 14.29 | TPCN2 |
| GO:BP | GO:0051126 | negative regulation of actin nucleation | 3.28E-02 | 12.5 | CTNNA2 |
| GO:MF | GO:0015085 | calcium ion transmembrane transporter activity | 3.67E-02 | 1.82 | CACNA1D |
| GO:BP | GO:0086012 | membrane depolarization during cardiac muscle cell action potential | 3.76E-02 | 5.88 | CACNA1D |
| GO:BP | GO:0086002 | cardiac muscle cell action potential involved in contraction | 3.76E-02 | 3.23 | CACNA1D |
| GO:BP | GO:0045761 | regulation of adenylate cyclase activity | 3.76E-02 | 4.55 | CACNA1D |
| GO:BP | GO:0070509 | calcium ion import | 3.76E-02 | 3.57 | CACNA1D |

| | | | | | |
|---|---|---|---|---|---|
| GO:BP | GO:0051349 | positive regulation of lyase activity | 3.76E-02 | 6.67 | CACNA1D |
| GO:BP | GO:0086001 | cardiac muscle cell action potential | 3.76E-02 | 3.33 | CACNA1D |
| GO:BP | GO:0086003 | cardiac muscle cell contraction | 3.76E-02 | 5.26 | CACNA1D |
| GO:BP | GO:0086010 | membrane depolarization during action potential | 3.76E-02 | 3.85 | CACNA1D |
| GO:BP | GO:0086091 | regulation of heart rate by cardiac conduction | 3.76E-02 | 2.56 | CACNA1D |
| GO:BP | GO:1904062 | regulation of cation transmembrane transport | 3.76E-02 | 3.33 | CACNA1D |
| GO:BP | GO:0031281 | positive regulation of cyclase activity | 3.76E-02 | 7.14 | CACNA1D |
| GO:BP | GO:0043266 | regulation of potassium ion transport | 3.76E-02 | 3.23 | CACNA1D |
| GO:BP | GO:0051928 | positive regulation of calcium ion transport | 3.76E-02 | 2.7 | CACNA1D |
| GO:BP | GO:0099623 | regulation of cardiac muscle cell membrane repolarization | 3.76E-02 | 4.55 | CACNA1D |
| GO:BP | GO:1901016 | regulation of potassium ion transmembrane transporter activity | 3.76E-02 | 3.85 | CACNA1D |
| GO:BP | GO:0032412 | regulation of ion transmembrane transporter activity | 3.76E-02 | 2.56 | CACNA1D |
| GO:BP | GO:0016338 | calcium-independent cell-cell adhesion via plasma membrane cell-adhesion molecules | 3.76E-02 | 5 | CLDN10 |
| GO:BP | GO:0051049 | regulation of transport | 3.76E-02 | 2.7 | CLDN10 |
| GO:BP | GO:0034315 | regulation of Arp2/3 complex-mediated actin nucleation | 3.76E-02 | 5.88 | CTNNA2 |
| GO:BP | GO:0048813 | dendrite morphogenesis | 3.76E-02 | 2.78 | CTNNA2 |
| GO:BP | GO:0050807 | regulation of synapse organization | 3.76E-02 | 3.85 | CTNNA2 |
| GO:BP | GO:0051147 | regulation of muscle cell differentiation | 3.76E-02 | 2.86 | CTNNA2 |
| GO:BP | GO:0120035 | regulation of plasma membrane bounded cell projection organization | 3.76E-02 | 3.57 | CTNNA2 |
| GO:BP | GO:2001222 | regulation of neuron migration | 3.76E-02 | 3.57 | CTNNA2 |
| GO:BP | GO:0048854 | brain morphogenesis | 3.76E-02 | 8.33 | CTNNA2 |
| GO:BP | GO:0051149 | positive regulation of muscle cell differentiation | 3.76E-02 | 3.7 | CTNNA2 |
| GO:BP | GO:0006939 | smooth muscle contraction | 3.76E-02 | 7.14 | TPCN2 |
| GO:BP | GO:0007040 | lysosome organization | 3.76E-02 | 2.63 | TPCN2 |
| GO:BP | GO:0080171 | lytic vacuole organization | 3.76E-02 | 3.03 | TPCN2 |
| GO:BP | GO:0043270 | positive regulation of ion transport | 4.68E-02 | 2 | CACNA1D |
| GO:BP | GO:1901379 | regulation of potassium ion | 4.81E-02 | 1.85 | CACNA1D |

| | | | | | |
|---|---|---|---|---|---|
| | | transmembrane transport | | | |
| GO:BP | GO:0051924 | regulation of calcium ion transport | 4.81E-02 | 1.72 | CACNA1D |
| GO:BP | GO:0009451 | RNA modification | 4.81E-02 | 1.72 | CDKAL1 |
| GO:BP | GO:0070830 | bicellular tight junction assembly | 4.81E-02 | 1.89 | CLDN10 |
| GO:BP | GO:0120192 | tight junction assembly | 4.81E-02 | 1.79 | CLDN10 |
| GO:BP | GO:0006400 | tRNA modification | 4.93E-02 | 1.54 | CDKAL1 |
| GO:BP | GO:0008033 | tRNA processing | 4.93E-02 | 1.56 | CDKAL1 |
| GO:BP | GO:0043297 | apical junction assembly | 4.86E-02 | 1.67 | CLDN10 |
| GO:BP | GO:0031329 | regulation of cellular catabolic process | 4.93E-02 | 1.54 | TPCN2 |

Supplementary Table 5. FLAME results of rare control SNPs with *gProfiler*.

| Source | Term_ID | Function | P-value | Enrichment Score % | Positive Hits |
|---|---|---|---|---|---|
| GO:CC | GO:1990350 | glucose transporter complex | 2.09E-02 | 100 | SLC2A1 |
| GO:BP | GO:1905204 | negative regulation of connective tissue replacement | 2.75E-02 | 50 | ABI3BP |
| GO:BP | GO:1905203 | regulation of connective tissue replacement | 2.75E-02 | 16.67 | ABI3BP |
| GO:BP | GO:1904597 | negative regulation of connective tissue replacement involved in inflammatory response wound healing | 2.75E-02 | 50 | ABI3BP |
| GO:BP | GO:1904596 | regulation of connective tissue replacement involved in inflammatory response wound healing | 2.75E-02 | 50 | ABI3BP |
| GO:BP | GO:0002248 | connective tissue replacement involved in inflammatory response wound healing | 2.75E-02 | 16.67 | ABI3BP |
| GO:BP | GO:1900454 | positive regulation of long-term synaptic depression | 2.75E-02 | 14.29 | IQSEC2 |
| GO:BP | GO:0050916 | sensory perception of sweet taste | 2.75E-02 | 14.29 | RGS21 |
| GO:BP | GO:0050917 | sensory perception of umami taste | 2.75E-02 | 14.29 | RGS21 |
| GO:BP | GO:0140271 | hexose import across plasma membrane | 2.75E-02 | 16.67 | SLC2A1 |
| GO:BP | GO:1904016 | response to Thyroglobulin triiodothyronine | 2.75E-02 | 20 | SLC2A1 |
| GO:BP | GO:0098708 | glucose import across plasma membrane | 2.75E-02 | 20 | SLC2A1 |
| GO:BP | GO:0098704 | carbohydrate import across plasma membrane | 2.75E-02 | 16.67 | SLC2A1 |
| GO:BP | GO:0002246 | wound healing involved in inflammatory response | 2.90E-02 | 12.5 | ABI3BP |

| | | | | | |
|---|---|---|---|---|---|
| GO:BP | GO:0097709 | connective tissue replacement | 2.98E-02 | 8.33 | ABI3BP |
| GO:BP | GO:0098696 | regulation of neurotransmitter receptor localization to postsynaptic specialization membrane | 2.98E-02 | 8.33 | IQSEC2 |
| GO:BP | GO:1901334 | lactone metabolic process | 2.98E-02 | 9.09 | SLC2A1 |
| GO:BP | GO:0070837 | dehydroascorbic acid transport | 2.98E-02 | 11.11 | SLC2A1 |
| GO:BP | GO:0015911 | long-chain fatty acid import across plasma membrane | 2.98E-02 | 10 | SLC2A1 |
| GO:BP | GO:0019852 | L-ascorbic acid metabolic process | 2.98E-02 | 9.09 | SLC2A1 |
| GO:BP | GO:1905209 | positive regulation of cardiocyte differentiation | 3.06E-02 | 7.69 | ABI3BP |
| GO:BP | GO:1900452 | regulation of long-term synaptic depression | 3.37E-02 | 6.67 | IQSEC2 |
| GO:BP | GO:0034104 | negative regulation of tissue remodeling | 3.61E-02 | 4.35 | ABI3BP |
| GO:BP | GO:1902473 | regulation of protein localization to synapse | 3.61E-02 | 4.35 | IQSEC2 |
| GO:BP | GO:1902683 | regulation of receptor localization to synapse | 3.61E-02 | 4.55 | IQSEC2 |
| GO:BP | GO:0032011 | ARF protein signal transduction | 3.61E-02 | 4.55 | IQSEC2 |
| GO:BP | GO:0032012 | regulation of ARF protein signal transduction | 3.61E-02 | 4.55 | IQSEC2 |
| GO:BP | GO:0044539 | long-chain fatty acid import into cell | 3.61E-02 | 5.88 | SLC2A1 |
| GO:BP | GO:0140354 | lipid import into cell | 3.61E-02 | 5 | SLC2A1 |
| GO:BP | GO:0071474 | cellular hyperosmotic response | 3.61E-02 | 5 | SLC2A1 |
| GO:BP | GO:1902001 | fatty acid transmembrane transport | 3.61E-02 | 5 | SLC2A1 |
| GO:BP | GO:1905207 | regulation of cardiocyte differentiation | 3.77E-02 | 4 | ABI3BP |
| GO:BP | GO:0090594 | inflammatory response to wounding | 3.77E-02 | 3.85 | ABI3BP |
| GO:BP | GO:0099633 | protein localization to postsynaptic specialization membrane | 3.77E-02 | 3.57 | IQSEC2 |
| GO:BP | GO:0099645 | neurotransmitter receptor localization to postsynaptic specialization membrane | 3.77E-02 | 3.57 | IQSEC2 |
| GO:BP | GO:0006972 | hyperosmotic response | 3.77E-02 | 3.7 | SLC2A1 |
| GO:BP | GO:0060292 | long-term synaptic depression | 4.31E-02 | 3.03 | IQSEC2 |
| GO:BP | GO:0051968 | positive regulation of synaptic transmission, glutamatergic | 4.58E-02 | 2.78 | IQSEC2 |
| KEGG | map04066 | HIF-1 signaling pathway | 4.72E-02 | 0.92 | SLC2A1 |
| KEGG | map04931 | Insulin resistance | 4.72E-02 | 0.93 | SLC2A1 |
| KEGG | map04911 | Insulin secretion | 4.72E-02 | 1.18 | SLC2A1 |

| | | | | | |
|---|---|---|---|---|---|
| KEGG | map04920 | Adipocytokine signaling pathway | 4.72E-02 | 1.43 | SLC2A1 |
| KEGG | map04919 | Thyroid hormone signaling pathway | 4.72E-02 | 0.83 | SLC2A1 |
| KEGG | map04976 | Bile secretion | 4.72E-02 | 1.12 | SLC2A1 |
| KEGG | map04922 | Glucagon signaling pathway | 4.72E-02 | 0.93 | SLC2A1 |
| KEGG | map05230 | Central carbon metabolism in cancer | 4.72E-02 | 1.43 | SLC2A1 |
| KEGG | map05211 | Renal cell carcinoma | 4.72E-02 | 1.47 | SLC2A1 |

Supplementary Table 6. FLAME results of rare control SNPs with *enrichR*

| Source | Term_ID | Function | P-value | Enrichment Score % | Positive Hits |
|---|---|---|---|---|---|
| GO:MF | GO:0015149 | hexose transmembrane transporter activity | 1.05E-02 | 6.67 | SLC2A1 |
| GO:MF | GO:0005355 | glucose transmembrane transporter activity | 1.05E-02 | 4.76 | SLC2A1 |
| GO:BP | GO:0046351 | disaccharide biosynthetic process | 1.25E-02 | 20 | SLC2A1 |
| GO:BP | GO:0098708 | glucose import across plasma membrane | 1.25E-02 | 20 | SLC2A1 |
| GO:BP | GO:0140271 | hexose import across plasma membrane | 1.25E-02 | 16.67 | SLC2A1 |
| GO:BP | GO:0019852 | L-ascorbic acid metabolic process | 1.25E-02 | 11.11 | SLC2A1 |
| GO:BP | GO:0044539 | long-chain fatty acid import into cell | 1.25E-02 | 10 | SLC2A1 |
| GO:BP | GO:0046323 | glucose import | 1.35E-02 | 7.69 | SLC2A1 |
| GO:BP | GO:0010827 | regulation of glucose transmembrane transport | 1.39E-02 | 5.26 | SLC2A1 |
| GO:BP | GO:1902001 | fatty acid transmembrane transport | 1.39E-02 | 5.26 | SLC2A1 |
| GO:BP | GO:1904659 | glucose transmembrane transport | 1.39E-02 | 5 | SLC2A1 |
| GO:BP | GO:0008645 | hexose transmembrane transport | 1.43E-02 | 4.35 | SLC2A1 |
| GO:BP | GO:0099177 | regulation of trans-synaptic signaling | 1.82E-02 | 2.86 | IQSEC2 |
| GO:BP | GO:0034762 | regulation of transmembrane transport | 1.82E-02 | 2.94 | SLC2A1 |
| GO:BP | GO:0019752 | carboxylic acid metabolic process | 1.92E-02 | 2.5 | SLC2A1 |
| GO:BP | GO:0090276 | regulation of peptide hormone secretion | 3.13E-02 | 1.35 | SLC2A1 |
| GO:BP | GO:0006767 | water-soluble vitamin metabolic process | 3.13E-02 | 1.32 | SLC2A1 |
| GO:BP | GO:0010232 | vascular transport | 3.13E-02 | 1.19 | SLC2A1 |
| GO:BP | GO:0150104 | transport across blood-brain barrier | 3.13E-02 | 1.16 | SLC2A1 |

| | | | | | |
|---|---|---|---|---|---|
| GO:BP | GO:0050804 | modulation of chemical synaptic transmission | 3.46E-02 | 0.92 | IQSEC2 |
| GO:BP | GO:0050796 | regulation of insulin secretion | 3.46E-02 | 0.96 | SLC2A1 |
| GO:BP | GO:0043933 | protein-containing complex subunit organization | 3.46E-02 | 0.89 | SLC2A1 |
| GO:BP | GO:0050708 | regulation of protein secretion | 3.67E-02 | 0.8 | SLC2A1 |
| KEGG | map05230 | Central carbon metabolism in cancer | 3.89E-02 | 1.49 | SLC2A1 |
| KEGG | map04976 | Bile secretion | 3.89E-02 | 1.41 | SLC2A1 |
| KEGG | map04922 | Glucagon signaling pathway | 3.89E-02 | 0.99 | SLC2A1 |
| KEGG | map05211 | Renal cell carcinoma | 3.89E-02 | 1.52 | SLC2A1 |
| KEGG | map04920 | Adipocytokine signaling pathway | 3.89E-02 | 1.43 | SLC2A1 |
| KEGG | map04931 | Insulin resistance | 3.89E-02 | 0.92 | SLC2A1 |
| KEGG | map04911 | Insulin secretion | 3.89E-02 | 1.18 | SLC2A1 |
| KEGG | map04066 | HIF-1 signaling pathway | 3.89E-02 | 0.97 | SLC2A1 |
| KEGG | map04919 | Thyroid hormone signaling pathway | 3.89E-02 | 0.85 | SLC2A1 |
| GO:CC | GO:0030864 | cortical actin cytoskeleton | 4.33E-02 | 2.38 | SLC2A1 |
| GO:CC | GO:0042383 | sarcolemma | 4.33E-02 | 1.92 | SLC2A1 |
| GO:CC | GO:0030863 | cortical cytoskeleton | 4.33E-02 | 1.72 | SLC2A1 |
| GO:MF | GO:0005085 | guanyl-nucleotide exchange factor activity | 4.89E-02 | 0.67 | IQSEC2 |
